# Whole-genome sequencing of half-a-million UK Biobank participants

**DOI:** 10.1101/2023.12.06.23299426

**Authors:** Shuwei Li, Keren J Carss, Bjarni V Halldorsson, Adrian Cortes, UK Biobank Whole-Genome Sequencing Consortium

## Abstract

Whole-genome sequencing (WGS) provides a comprehensive view of the genome, enabling detection of coding and non-coding genetic variation, and surveying complex regions which are difficult to genotype. Here, we report on whole-genome sequencing of 490,640 UK Biobank participants, building on previous genotyping^1^ and whole-exome sequencing (WES) efforts^2,3^. This advance deepens our understanding of how genetics influences disease biology and further enhances the value of this open resource for the study of human biology and health. Coupling this dataset with rich phenotypic data, we surveyed within- and cross-ancestry genomic associations with health-related phenotypes and identified novel genetic and clinical insights. While most genome-wide significant associations with disease traits were primarily observed in Europeans, we also identified strong or novel signals in individuals of African and Asian ancestries. Deeper capture of exonic variation in both coding and UTR sequences, strengthened and surfaced novel insights relative to WES analyses. This landmark dataset, representing the largest collection of WGS and available to the UK Biobank research community, will enable advances into our understanding of the human genome, and facilitate the discovery of new diagnostics, therapeutics with higher efficacy and improved safety profile, and enable precision medicine strategies with the potential to improve global health.

Graphic summary.
Framework of the WGS UKB study. This figure captures the flow of this manuscript. We start with the collection of patient samples by UK Biobank and followed by the strategy taken to perform WGS. We continue with quality control performed on GraphTyper and DRAGEN datasets, followed by variant calling of SNPs, in/dels, and structural variants (SV). Thereafter we defined the phenotypes (binary and quantitative) associated with SV, SNPs and at the gene level (rare variant analysis) and conclude with the definition of five ancestry groups and collective association effect as a cross-ancestry meta-analysis.

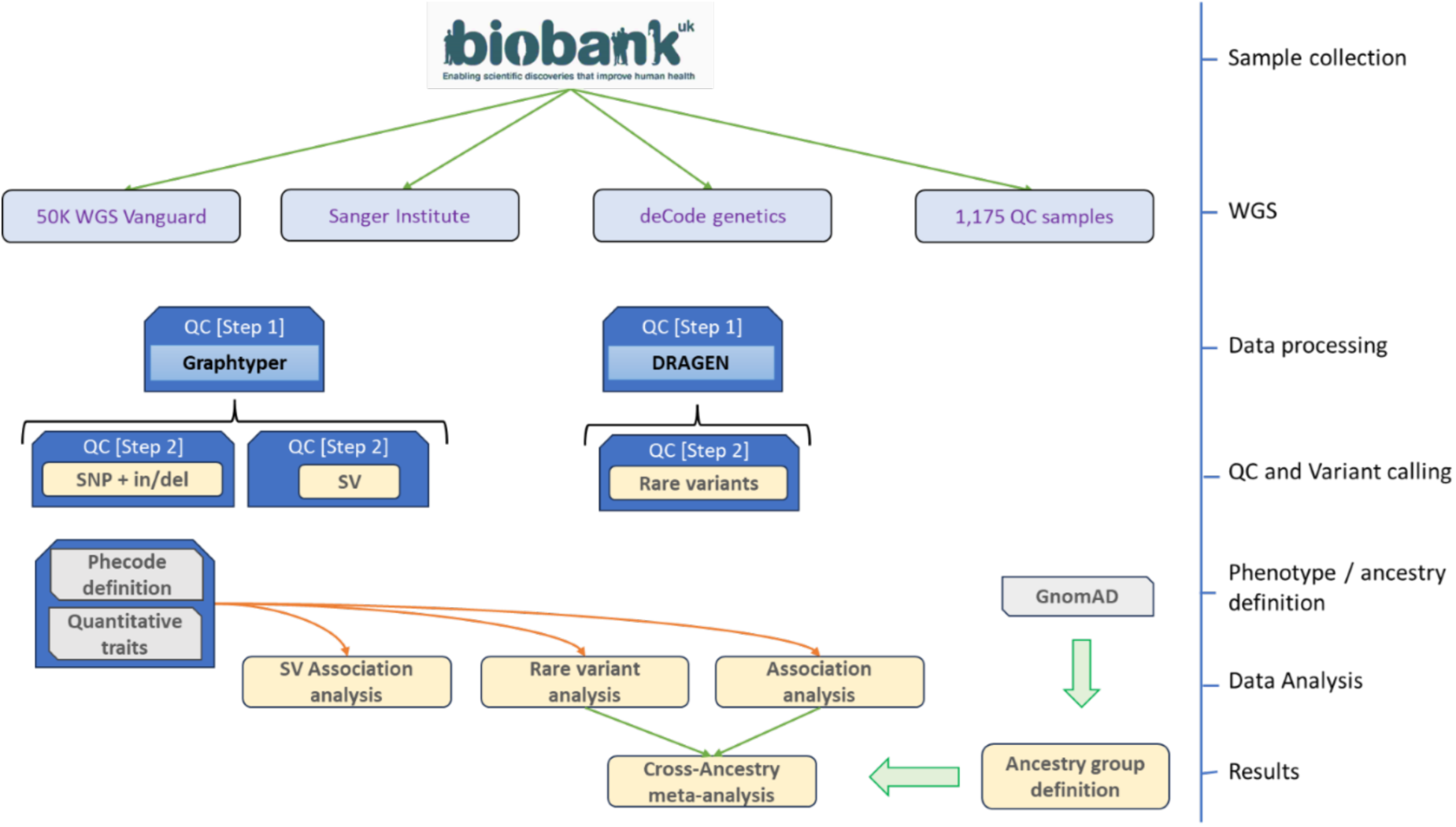

## Main Text

The UK Biobank (UKB) is a population-based study that collected detailed information from half a million UK participants, including biological samples and comprehensive health-related and demographic measures^1^. Numerous subsequent data collection and generation efforts, including multimodal brain imaging^4^, proteomics^5^, metabolomics^6^ and others, have dramatically increased the depth of the dataset. Here, we present a step change in the UK Biobank resource, and for the life sciences, with the completion of whole genome sequencing in half a million participants. In the original release, all samples were genotyped^1^ and imputed to ∼96 million single nucleotide polymorphisms (SNP). SNP genotyping and imputation allow the accurate characterization of relatively common variants, but it is inaccurate for rare-genetic variation and complex regions of the genome. UKB samples also underwent whole exome sequencing^7^ (WES), which allows for characterization of the 2-3% exonic portion of the genome but omits nearly all non-coding variation and is limited in the detection of structural variants. Rare non-coding variation is known to contribute to human diseases and complex traits, although this remains relatively understudied^8–10^. This large-scale, deeply phenotyped WGS dataset brings enormous potential to expand our understanding of the role of rare non-coding variation in health and disease.

In this report, we demonstrate the utility of WGS in the identification of ∼1.5 billion variants (comprising of SNP, indel variants and structural variants) in the UK Biobank participants. We observed an 18.8-fold and over 40-fold increase in observed human variation compared to imputed array and WES, respectively. These variants were associated with many disease features and traits, enabling improved characterization of disease mechanisms, such as variants influencing disease risk via non-coding mechanisms. These data can be used to address multiple drug discovery and development questions, including target selection, validation, assessment of safety concerns, identification of patient populations with specific underlying genetic drivers of disease, and repositioning opportunities ^11,12^. A valuable unique benefit is that these data will facilitate an improved understanding of the selective constraints acting on disruption outside the coding genome which will improve the ability to prioritize rare non-coding variants with a large effect on disease risk^13^.

This unprecedented resource will greatly enable exploration of human genetic variation and its implication on disease etiology. Here we describe the resource and highlight some initial examples of unique insights and future avenues for exploration.

## Results

### Data processing

#### Sequencing

The whole genomes of 490,640 UKB participants were sequenced to an average coverage of 32.5x (> 23.5x per individual, Supplementary Fig. S1) using Illumina NovaSeq 6000 sequencing machines, in addition, 1,175 samples were sequenced in duplicate for quality control (QC) purposes (see Supplementary Methods).

#### Cohorts

We defined five cohorts with distinct ancestry in the UK Biobank WGS dataset using a classifier trained with data from the Genome Aggregation Database^16^ (gnomAD) (Methods), which identified 9,229 participants being of African ancestry (AFR), 2,869 of Ashkenazi ancestry (ASJ), 2,245 of East Asian ancestry (EAS), 458,855 of non-Finnish European ancestry (NFE) and 9,674 of South Asian (SAS) ancestry, and remaining 7,768 individuals of other ancestries or non-confidently assigned to one group. Most individuals (93.5%) were of non-Finnish European ancestry, with the remaining 31,785 individuals representing other continental populations. While this resource is largely European, this effort marks the largest WGS effort to date in non-European individuals. The relative increment is largest in the South Asian ancestry group, where the UKB WGS SAS cohort is four times larger than any other WGS cohort of this ancestry available in the Genome Aggregation Database (gnomAD v3; 2,419 SAS individuals), the 1000 Genomes Project (1KPG; 601) or the Human Genome Diversity Project (HGDP; 181).

#### SNP and indels

This study reports findings from three different SNP and indel datasets: 1) joint calling across all individuals using GraphTyper, 2) single sample calling with DRAGEN 3.7.8, and 3) multi-sample aggregated DRAGEN 3.7.8 dataset (Methods). This diversity of approaches reflects developments to these methods throughout the course of this project and gives the opportunity to explore the various workflows used by consortium members and other users of the UKB. We expect the different datasets to yield highly comparable results, however systematic comparison of the strengths and weaknesses of different methods is outside the scope of this manuscript.

We called 1,037,556,156 SNPs and 101,188,713 indels using GraphTyper (Figure 1a). A large majority of variants, 1,025,188,151 (98.80%) SNPs and 97,190,353 (96.05%) indels were reliable (AAscore > 0.5 and < 5 duplicate inconsistencies; Methods). All GraphTyper analyses are restricted to this set unless otherwise noted. The number of variants identified per individual using GraphTyper was, on average, 43 times larger than the number of variants identified through WES ^17^ (Table 1a, Methods). Notably WES misses variants in exons that are transcribed but not translated, 69.2% and 89.9% of the 5’ and 3’ untranslated region (UTR) variants are missing from WES, respectively. Even inside of coding exons currently curated by Encode^18^, we estimate that 13.7% of variants are missed by WES (Table 1a, Table S1, Table S2). A subset of the missed variants is explained by the 25,853 fewer samples that are available in the WES release. Manual inspection of a subset of the missing variants in WES, where both WES and WGS were available, suggests these are absent due to both missing coverage in some regions as well as genotyping filters. Almost all variants identified with WES are found by WGS (Table 1a).

**Figure 1.**
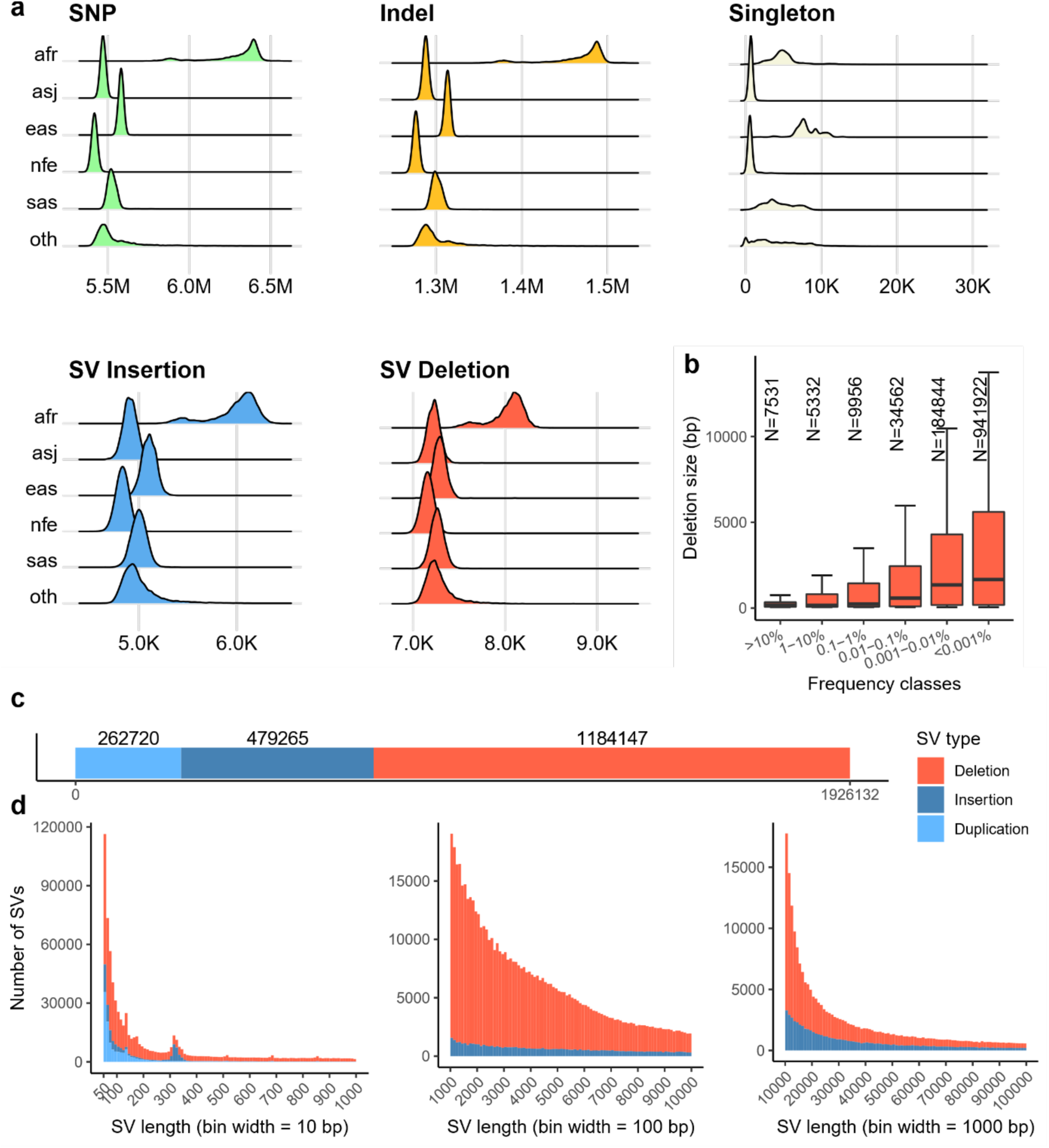
a) Panels showing the density/counts of the per-individual number of variants split up by the five populations considered in this study. Figures show number of SNPs, indels, singleton SNPs and indels, combined number of SV insertions and duplications and SV deletions. b) The length of SV deletions discovered in this study, split by the frequency of the variant. Red area shows the size of variants in 25th-75th quartile. Middle line shows the median length and top horizontal line the 95th percentile. c) The number of variants discovered split by variant class, duplication, insertion and deletion. d) The size of insertions and deletions discovered shown in range from 50bp up to 1,000, 10,000 and 100,000 bp.

**Table 1.**
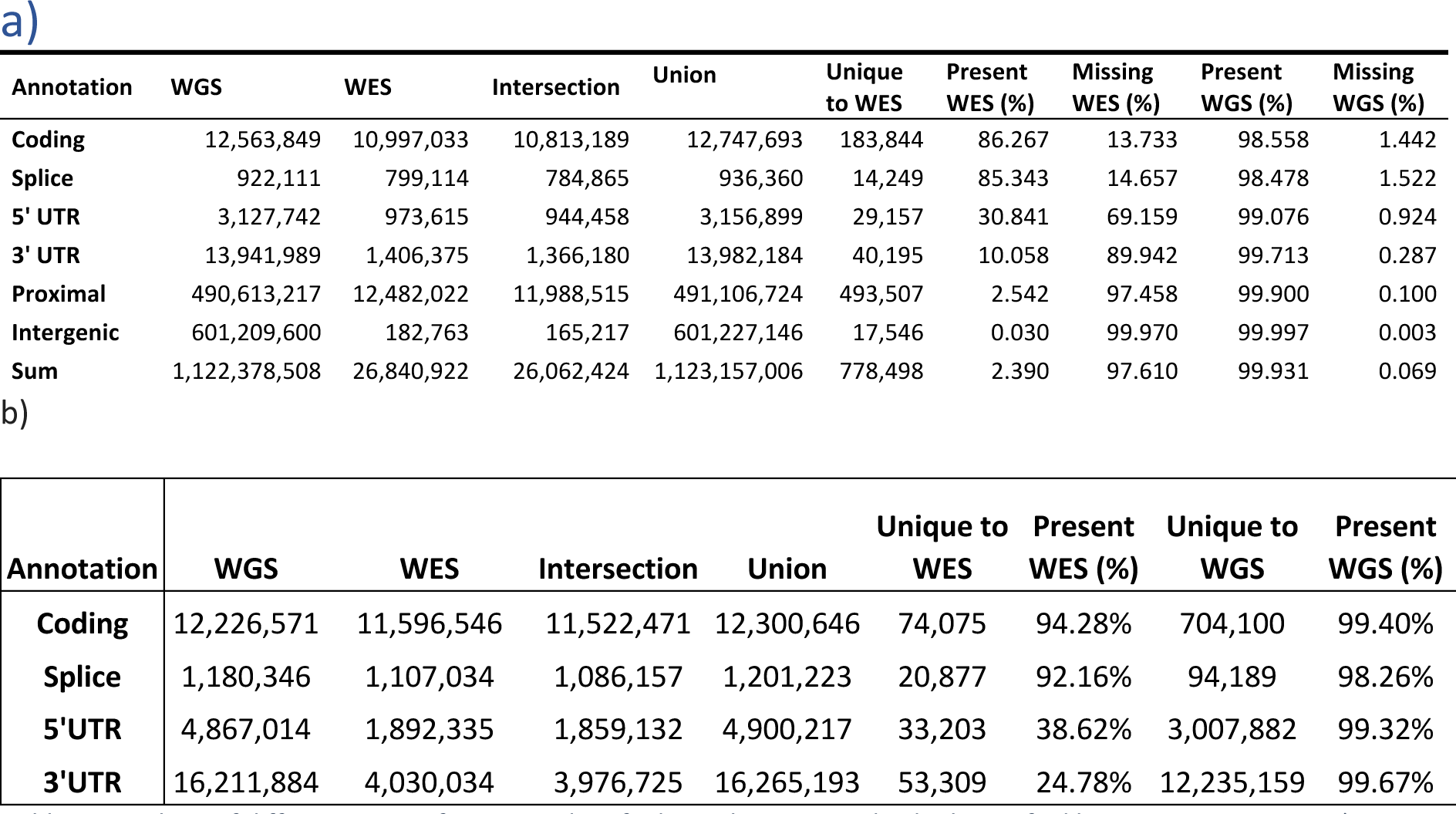
Numbers of different types of variants identified in at least one individual stratified by annotation across a) GraphTyper dataset, using Ensembl version 101 annotations comparing WES and WGS data releases and b) DRAGEN single sample dataset annotated using SnpEff v4.3 against Ensembl Build 38.92. For DRAGEN, high quality variant counts are limited to the 460,552 samples for whom we had both WES and WGS available, and percentages are based on the number of variants compared to the union across WES and WGS variants per annotation type. Table S3 provides the DRAGEN aggregated dataset variant counts across the extended list of variant classes and for all WGS participants.

| Annotation | WGS | WES | Intersection | Union | Unique to WES | Present WES (%) | Missing WES (%) | Present WGS (%) | Missing WGS (%) |
| --- | --- | --- | --- | --- | --- | --- | --- | --- | --- |
| **Coding** | 12,563,849 | 10,997,033 | 10,813,189 | 12,747,693 | 183,844 | 86.267 | 13.733 | 98.558 | 1.442 |
| **Splice** | 922,111 | 799,114 | 784,865 | 936,360 | 14,249 | 85.343 | 14.657 | 98.478 | 1.522 |
| **5' UTR** | 3,127,742 | 973,615 | 944,458 | 3,156,899 | 29,157 | 30.841 | 69.159 | 99.076 | 0.924 |
| **3' UTR** | 13,941,989 | 1,406,375 | 1,366,180 | 13,982,184 | 40,195 | 10.058 | 89.942 | 99.713 | 0.287 |
| **Proximal** | 490,613,217 | 12,482,022 | 11,988,515 | 491,106,724 | 493,507 | 2.542 | 97.458 | 99.900 | 0.100 |
| **Intergenic** | 601,209,600 | 182,763 | 165,217 | 601,227,146 | 17,546 | 0.030 | 99.970 | 99.997 | 0.003 |
| **Sum** | 1,122,378,508 | 26,840,922 | 26,062,424 | 1,123,157,006 | 778,498 | 2.390 | 97.610 | 99.931 | 0.069 |

Table 1. Numbers of different types of variants identified in at least one individual stratified by annotation across a) GraphTyper dataset, using Ensembl version 101 annotations comparing WES and WGS data releases and b) DRAGEN single sample dataset annotated using SnpEff v4.3 against Ensembl Build 38.92.
| Annotation | WGS | WES | Intersection | Union | Unique to WES | Present WES (%) | Unique to WGS | Present WGS (%) |
| --- | --- | --- | --- | --- | --- | --- | --- | --- |
| **Coding** | 12,226,571 | 11,596,546 | 11,522,471 | 12,300,646 | 74,075 | 94.28% | 704,100 | 99.40% |
| **Splice** | 1,180,346 | 1,107,034 | 1,086,157 | 1,201,223 | 20,877 | 92.16% | 94,189 | 98.26% |
| **5'UTR** | 4,867,014 | 1,892,335 | 1,859,132 | 4,900,217 | 33,203 | 38.62% | 3,007,882 | 99.32% |
| **3'UTR** | 16,211,884 | 4,030,034 | 3,976,725 | 16,265,193 | 53,309 | 24.78% | 12,235,159 | 99.67% |

We compared the DRAGEN single sample WGS dataset to the previously published DRAGEN WES dataset^19^ to explore the number of variants identified across coding, splice and 5ʹ and 3ʹ untranslated region (UTR) annotation categories. As previously described^15^ a greater number of variants were captured in the WGS data across all annotation categories, with the majority (98.26%-99.67%) of variants identified in the WES dataset being captured in the WGS data (Table 1b). WES did not capture many of the UTR variants, particularly 3’UTR variants where only 24.78% of variants present across both datasets were found in the WES data, compared to 99.67% in the WGS data (Table 1b). Reassuringly, the pattern of variant-numbers was generally similar between GraphTyper and DRAGEN single sample datasets.

Using the DRAGEN multi-sample dataset we called 1,289,650,789 SNPs and 204,960,409 indels on autosomes, sex chromosomes, mitochondria and alternate contigs of 490,541 individuals (Table S3, Table S4). After using DRAGEN Machine Learning Recalibration cutoff QUAL>=3 we assessed the sample-level variant calling accuracy using Genome in a Bottle samples and we achieve SNP sensitivity 99.77%, SNP precision 99.91%, indel sensitivity 99.70%, and indel precision 99.83% (Table S4), within Genome in a Bottle high-confidence regions. Variants are aggregated and genotyped using DRAGEN Iterative gVCF Genotyper with the same quality cutoff to maintain variant accuracy, which yielded 1,109,854,569 variant sites. We next evaluated the genotyping at these variant sites and found 1,010,107,317 (91%) have genotyping rate above 90%, comprising 1,108,231,808 SNPs and 148,001,752 indels (see Methods and Table S5). Using random downsampling of samples, we investigated the gain in number of variants in the UK Biobank DRAGEN aggregated variant dataset as sample size increases from 1000 to 490,541 (Figure 2). As expected, for common variants (e.g., >1% frequency) we observe only modest increase in number of variants with increasing sample size, but for the rarest variants (e.g., <=0.001% frequency), we observe substantial increases in number of variants with sample size, that don’t appear to reach a plateau even at the highest sample size, supporting the value of continuing very large-scale sequencing efforts to discover novel and high-impact rare variants (Figure 2).

**Figure 2.**
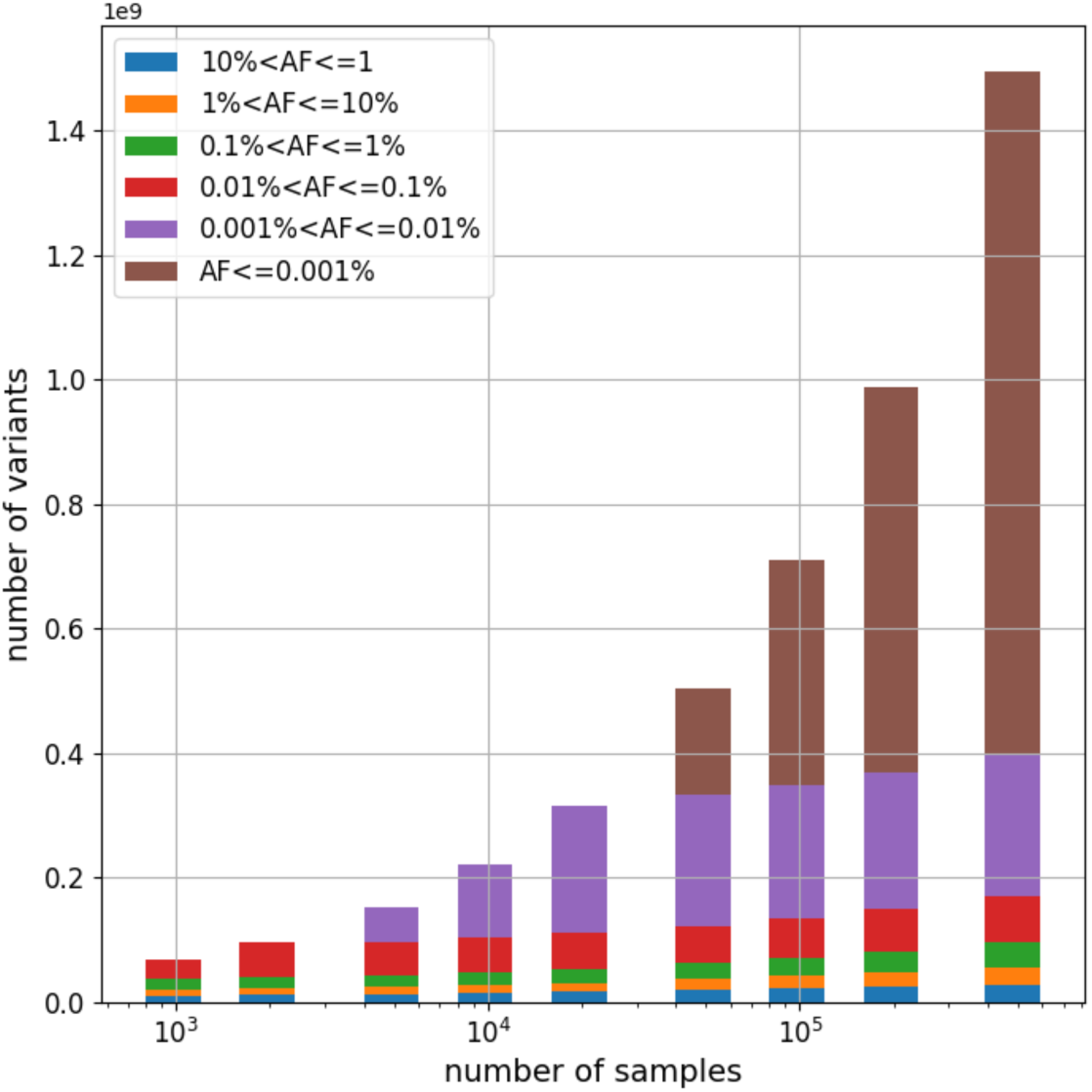
Number of variants in UK Biobank DRAGEN aggregated variant dataset in different allele frequency ranges as the number of samples increase from 1000 to 490,541 (based on random downsampling). Variant alleles are collected from all autosomes, sex chromosomes, mitochondria, and ALT contigs.

#### Structural variants

We identified structural variants (SVs) in each individual using the DRAGEN SV caller and combined these with variants from a long-read study^20^ and the assemblies of seven individuals^21^. The resulting 2,739,152 SVs were genotyped with GraphTyper^21^, of which 70.3% (1,926,132) (Figure 1b) were considered reliable; 262,720 duplications, 479,265 insertions and 1,184,147 deletions. SVs were defined as variants being at least 50bp and size distribution showed a well-documented skew toward short variants (Figure 1b).

On average we identified 13,102 reliably called SVs per individual, 7,340 deletions and 5,762 insertions or duplications (Figure 1a). These numbers are greater than the 7,439 SVs per individual found by gnomAD-SV^22^, another short-read study, but considerably smaller than the 22,636 high quality SVs found in a long-read sequencing study^20^ mostly due to an under-representation of insertions and SVs in repetitive regions. Despite the number of SVs being much smaller than the number of SNPs and indels, the number of base pairs impacted per haploid genome on average (3.6 Mbp) is comparable to that of SNPs (2.9 Mbp) and indels (1.5 Mbp). Most of the structural variants are very rare; 1,470,329 (76.3%) are carried by fewer than 10 individuals (<0.001% frequency). We observed that rare variants are generally longer than common variants with a median length of 1,660 bp for deletions carried by fewer than 10 individuals and 169 bp for deletions with frequency above 1% (Figure 1b).

Variant identification was performed analogously to the UKB 150k release^15^ but replacing Manta^23^ with the DRAGEN SV caller which identifies a greater number of insertions. Due to the improved discovery step and a modified variant filtering procedure the number reliably called SVs is approximately 3-fold larger in the current set compared to the previous release^15^. Out of the 637,321 SVs reliably called in our previous call set, 590,037 (92.6%) are also reliably called in the current call set. An additional 11,958 (1.8%) were part of the genotyping set but no longer considered reliable when genotyped while the remaining 35,327 (5.5%) were not part of the current set of variants.

The number of variants called per individual varies by population, with the largest number of variants called in individuals in the AFR cohort, followed by the EAS, SAS, ASJ and finally NFE, where individuals had the fewest number of called variants when compared to the current reference genome which is mostly of European descent (Figure 1a).

#### Phenotype associations

We integrated deep phenotyping data^24^ available for the majority of UKB participants and performed genetic association analysis across selected disease outcomes captured with electronic health records and molecular and physical measurement phenotypes, many of which are well-established disease biomarkers. Association testing was performed in all observed genetic variants and using several genetic models, we included single variant tests, multi-ancestry meta-analysis, rare variant collapsing analysis, and structural variant analysis (Methods).

#### Genome-wide association analysis of the UK Biobank phenome

Genome-wide association analysis for individual SNPs and small indels was performed using the GraphTyper dataset in each ancestry cohort for 764 ICD-10 codes (N cases > 200) and 71 selected quantitative phenotypes (N > 1,000) (Table S6). For the NFE cohort, we estimated the gain in discovery and improvement of fine-mapping in association signals against variants observed in the imputed array genetic dataset^1^. We observed that whilst the increase in discovery was modest for common variant associations (Fig. S2), the ability to fine-map association signals was improved. We identified 33,123 associations across 763 binary and 71 quantitative GWAS datasets (Methods). Of these, 3,991 (12.05%) are novel to the WGS data when compared to those identified using only array imputed variants. As expected, the majority of associated variants novel to WGS are rare variants, including 86% of associations with MAF < 0.0001, while only 2% of associations with MAF > 0.1 are novel to WGS (Fig. S2). Among the 29,357 associations identified using array imputed variants, 2,984 had a different, more significant, lead variant in the WGS variants, resulting in improved fine-mapping of the association signals observed (Table S7). For example, a common variant association uncovered by WGS that was previously missed by the imputed array data is near genes *MRC1* and *TMEM236* in chromosome 10, where we identified an association between rs371858405 (NFE MAF = 0.24) and reduced hypothyroidism risk (OR = 0.94, p-value = 2.6e-11). In the imputed data, the region within the WGS lead-variant has sparse SNP coverage when compared to adjacent regions (Fig. S3a), likely a result of a patch to the hg19 reference genome (chr10_gl383543_fix) that occurred after the UK Biobank genotyping array was designed. A second example illustrating novel biological findings with rare genetic variation is the observation of a rare frameshift variant (MAF = 5.1 10^-5^) in *FOXE3* chr1:47417015:GC:G (rs1176723126) found to be significantly associated with the first occurrence phenotype “other cataract” (H26), p-value=6.2 10^-9^ (Fig. S3b). The link between *FOXE3* and cataract, and other ocular diseases, was reported in previous familial studies and human and mouse disease models^25^, but the association was not observed in the UKB imputed array and meta-analysis that included UKB imputed array^26^.

#### Multi-ancestry meta-GWAS

To examine multi-ancestry genetics of tested health-related phenotypes, we performed trans-ancestry meta-analysis of the GraphTyper GWAS data across five ancestries for 68 quantitative traits with ≥1,000 measurements in at least two ancestries and 228 ICD-10 disease outcomes with ≥200 cases in at least two ancestries. We identified 28,674 genome-wide significant (p<5.0e-08) sentinel associations in the meta-analysis (Methods, Table S8), including 1,934 meta-only significant associations, 26,478 associations significant only in NFE, and 82 nonNFE-only associations (Table S9; Figure 3).

**Figure 3.**
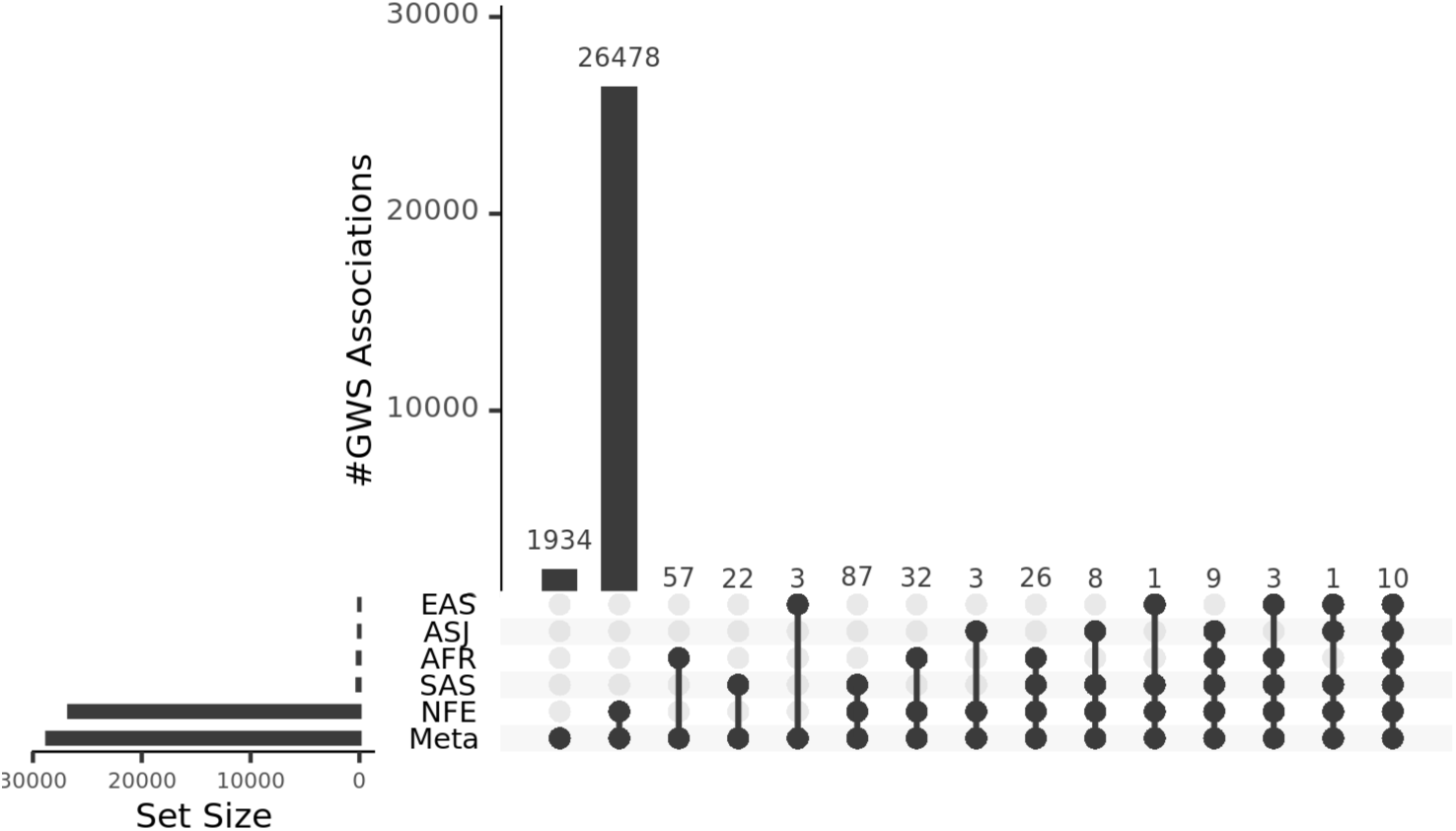
UpSet plot of GWS (genome-wide significant) associations across ancestries. Ancestry labels are sorted by #of GWS associations in each set: meta-analysis, NFE (non-Finnish European), SAS (South Asian), AFR (African), ASJ (Ashkenazi Jewish), EAS (East Asian).

For example, it is well-known that APOE e2 alleles (rs7412-T and rs429358-T) protect carriers against Alzheimer’s disease (AD)^27^, while e4 alleles (rs7412-C and rs429358-C) increase the AD risk. We found rs7412-T is significantly associated with lower Apolipoprotein B (apoB), Lipoprotein A, LDL, total cholesterol, and lower risk of lipoprotein metabolism disorder (ICD10: E78) across ancestries with concordant effects. This is consistent with published evidence of rs7412-T association with lipid biomarkers in Europeans, Africans, Asians, and Hispanics^28–36^; lower apoB has been linked to reduced AD risk, with causality to be confirmed^32,37–43^. As expected, rs429358-C, is associated with higher risk of dementia and cognitive disorders (F01-F09), intracranial hemorrhage and cerebrovascular disease (ICD10: I60-I69). However, rs429358-C is also associated with lower C-reactive protein (CRP) across ancestries and lower liver disease (ICD10: K70-K77) risk in meta-analysis from non-significant concordant signals in AFR and SAS. Further, it is associated with lower risk of obesity (ICD10: E66) and pulmonary disease (ICD10: J44) with meta-results dominated by NFE signals. Recent studies observed same effect of rs429358-C on CRP in Europeans, Japanese and Koreans, strongly suggesting higher circulating CRP increases the risk of age-related macular degeneration^34,44–47^ Prior studies also reported rs429358-C’s association with lower NFLD (nonalcoholic fatty liver) and liver enzyme level in Europeans, Africans, Asians and Hispanics^48–55^; but novel associations with obesity and pulmonary disease are detected here, with smaller effect size than other reported diseases and requires further investigation.

Of the meta-analysis significant associations, 126 were driven by non-NFE ancestries despite the much smaller sample size compared to NFE (Fig. S4a): 83 with strongest signals (sentinel variants with smallest p-value) in AFR, 37 in SAS, 5 in EAS and 1 in ASJ. Almost all the non-NFE driven significant sentinel variants (80 AFR, 37 SAS, 5 EAS and 1 ASJ) had MAF (minor allele frequency) < 0.5% in NFE; the median MAF enrichment compared with NFE is highest in AFR (MAF_AFR_/MAF_NFE_] = 828.49, followed by EAS and SAS with relative wide range of enrichment (Fig. S4b), suggesting these signals were driven by higher allelic enrichment shaped by population history. For example, the HBB-HBE1 locus’ association with Anemia (ICD10: D55-59) has the strongest signals in AFR and SAS, but no signals at all in NFE. The sentinel SNP rs334 (11-5227002-T-A, missense variant in HBB gene) is the most common cause of sickle cell disease (SCD), leading to abnormal hemoglobin (HbS) and sickle cell anemia. In this study, rs334-A is common in AFR (AF=6.3%) but rare (AF<0.08%) in NFE and SAS (Figure 4); similarly, from gnomAD (v4.0.0), rs334-A is common in Africans (AF=4.9%), rare in South Asian (AF=0.09%) and almost absent in Europeans (AF=0.003%). This is driven by rs334-A’s protective effect against malaria^56^, despite its pathogenic effect causing SCD, and carriers’ survival advantage under selection pressure in areas with high malaria prevalence (based on 2022 world malaria report, 95% malaria cases were in Africa, followed by 3-4% in South-East Asia; while the European region has been free of malaria since 2015). We observed rs334-A is also associated with higher Cystatin-C, blood urea and lower urinary creatine, sodium, potassium, which is expected as one severe complication of SCD is sickle cell nephropathy primarily attributed to hemolysis and vascular occlusion^57^. Another HBB nonsense variant rs11549407 (11-5226774-G-A) is the sentinel signal strongly associated (p-value<5.6e-62, beta=6.9) with beta Thalassaemia (ICD10: D56), despite its low frequency in NFE (AF=0.005%) and absence in other ancestries (Figure 4); from gnomAD, rs11549407-A has an AF of 0.02% in non-Finnish Europeans, 0.003% in Africans, completely absent in Asian and Ashkenazi Jewish. rs11549407-A introduces a premature stop codon, leading to a truncated beta-globin protein and reduced production of normal beta-globin chains, thus unstable hemoglobin molecules and Thalassemia. In contrast to rs334, this nonsense mutation with strong disease-causing effect has not been shown to confer protection against malaria or other pathogens. One other HBB splice site variant rs33915217 (11-5226925-C-G), which is also pathogenic for beta Thalassaemia (Figure 4), is associated with lower corpuscular haemoglobin concentration specifically in SAS (AF=0.4%) with no signals from other ancestries; from gnomAD, it also occurs more commonly in South Asian (AF=0.5%) and almost absent in Africans and Europeans. The higher frequency of this variant in SAS could be shaped by genetic drift, founder effect or potential selective advantage specific to South Asians, which is yet to be established given limited report on rs33915217^58^. Under the same selection pressure of malaria, another G6PD missense variant rs1050828 (X-154536002-C-T), which cause the G6PD deficiency and hemolytic anemia, but provides protection against severe malaria, reaching high frequency in AFR (14.7%) but is rare in NFE (0.005%). Therefore, despite the much larger NFE sample size, here we detected rs1050828-T as an AFR specific GWS signal, associated with higher reticulocyte count/percentage/volume and total bilirubin, attributing to compensatory release of more reticulocytes triggered by hemolysis.

**Figure 4.**
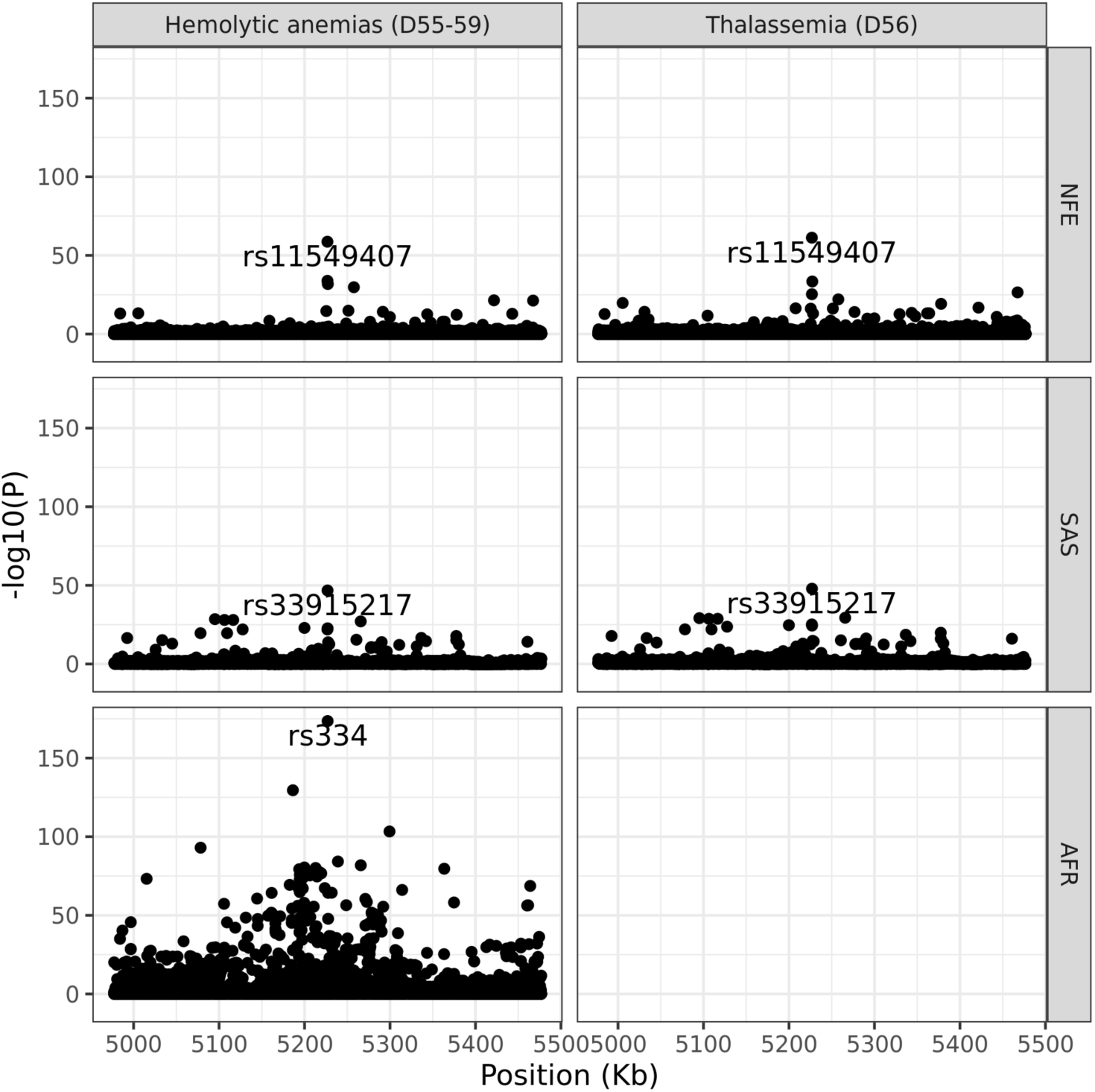
Regional plot for HBB-HBE1 locus associated with a) Hemolytic anemias. (D55-59) in NFE, AFR, SAS and **b) Thalassaemia** (D56) in NFE, AFR, SAS. NFE: non-Finnish European; AFR: African; SAS: South Asian; EAS: East Asian; ASJ: Ashkenazi Jewish.

#### Loss-of-function (LoF) variants in WGS

Naturally occurring human genetic variation known to result in disruption of protein-coding genes provide an *in vivo* model of human gene inactivation. Individuals with LoF (loss-of-function) variants, particularly those with homozygous genotypes, can therefore be considered a form of human “knock-outs” (KOs). Studying human KOs affords an opportunity to predict phenotypic consequences of pharmacological inhibition. Besides putative LoF (pLoF) variants that can be predicted based on variant annotation, ClinVar ^24^ also reported pathogenic/likely pathogenetic (P/LP) variants with clinical pathogenicity. Among the 490K UKB WGS samples (GraphTyper dataset), we found 10,071 autosomal genes with at least 100 heterozygous carriers of pLoF/P/LP variants and 1,202 autosomal genes with at least three homozygous carriers. Especially, among the 81 genes recommended by the ACMG^59^ (American College of Medical Genetics and Genomics) for clinical diagnostic reporting, we found 7,313 pLoF/P/LP variants carried by 51,107 individuals. Furthermore, there are 81 homozygous carriers of pLoF/P/LP found in 14 ACMG genes; of which 56 participants carry mutations in DNA repair pathway genes like MUTYH, PMS2, MSH6 (Table S10). Among them, a subset are clinically actionable genotypes with a confirmed functional impact in corresponding inheritance mode. Further validation, and confirmation with ACMG diagnostic criteria, is needed to determine which variants are clinically actionable.

Specifically, for *PCSK9*, WGS revealed 961 heterozygous carriers of pLoF/P/LP variants and one homozygous carrier of a LoF (rs28362286, p.C679X), which is relatively common in Africans but rare in Europeans (gnomAD AF: 0.8% vs. 0.004%). As with PCSK9 inhibitors, C679X has been shown to lower LDL levels ^60–64^. A homozygote carrier of C679X, who has African ancestry, exhibits remarkably low plasma LDL level (1.49 mmol/L, Fig. S5). Comparing UKB WGS vs. WES datasets, among the same set of 450K participants, ∼16000 autosomal genes harboring pLoF/P/LP variants in ≥1 carriers in both WGS and WES; but 1,977 more genes can be found in WGS with at least 100 carriers of pLoF/P/LP variants (Figure 5), this is expected given the WGS library design and deeper coverage.

**Figure 5:**
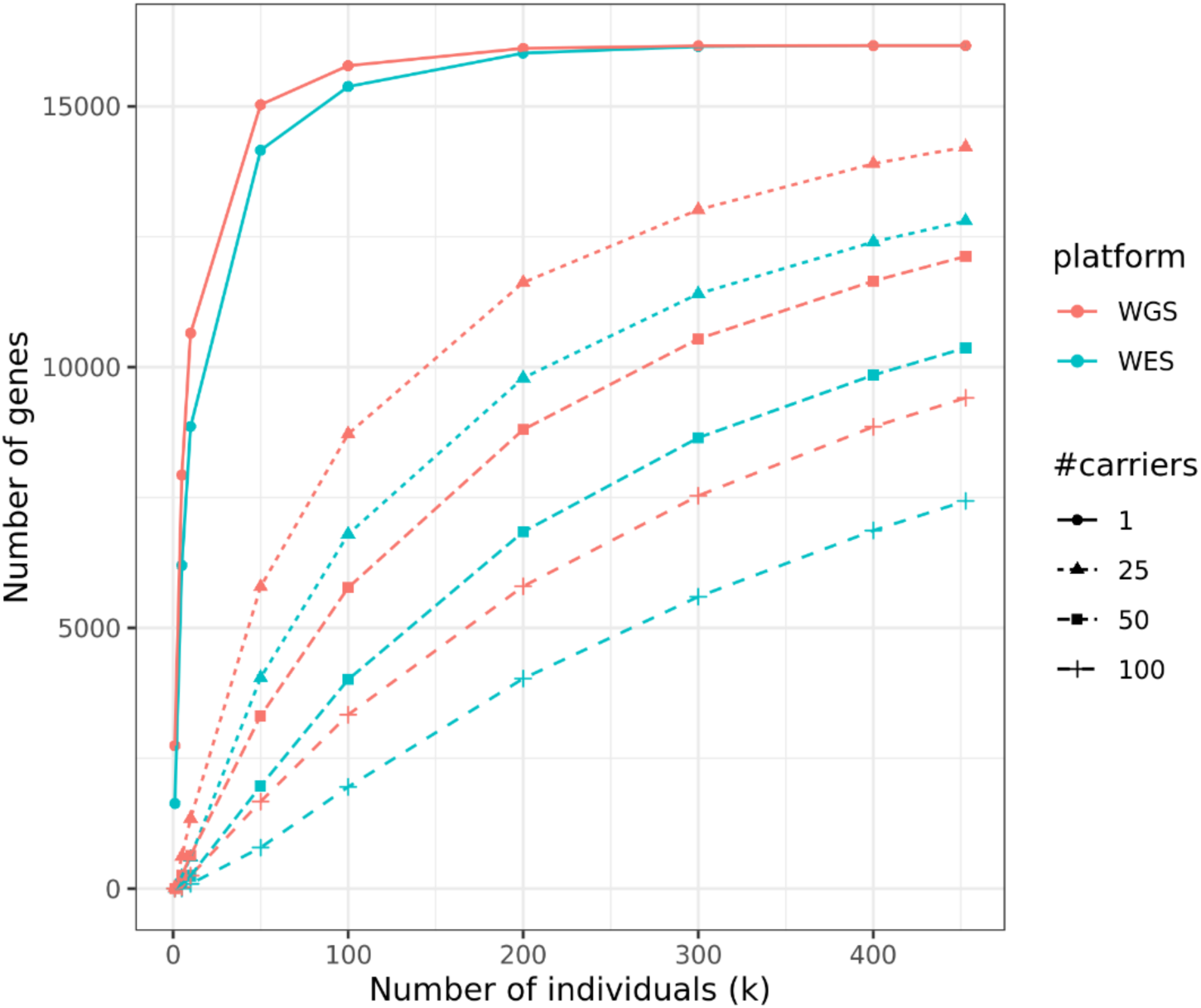
Observed number of genes in carriers of heterozygous pLoF/P/LP variants in WGS and WES. The number of autosomal genes (Y-axis) with at least 1, 25, 50 and 100 carriers of heterozygous pLoF variants among the number of individuals (X-axis) to the total number of 452,728 participants with both WES and WGS data. pLoF: putative loss-of-function variants. P/LP: ClinVar pathogenic/likely pathogenic variants (see Supplementary Methods for more details of pLoF and P/LP definitions).

#### Rare variant collapsing analyses PheWAS of exons in WGS compared to WES

All results for rare variant collapsing analyses use the single sample variant calls generated as part of the UKB DRAGEN-processed data releases. Gene-level collapsing analysis, in which aggregation of rare variants is tested for association with disease, has emerged as a powerful method for identifying gene-phenotype associations driven by high allelic heterogeneity^19,65^. To date, most collapsing analyses have used WES data^66^, requiring careful filtering for inadequately sequenced sites. We hypothesized that the greater coverage of WGS compared to WES, and improved sequence representation across certain technically challenging regions of the exome, could increase power to detect gene-phenotype associations in phenome-wide association studies (PheWAS). We therefore performed two separate collapsing analysis-based PheWAS on an identical sample of 460,552 individuals using both WES- and WGS-based coding regions processed by DRAGEN (Methods).

In total, we tested for the association between 18,930 genes and 751 phenotypes (687 binary “First Occurrence” phenotypes and 64 quantitative traits that met our inclusion criteria, see methods and Table S11) using 10 different non-synonymous collapsing analysis models plus a synonymous control model (Table S12) as previously described^19^ (Methods). We performed a multi-ancestry meta-analysis to estimate the aggregate effect of qualifying variants (QVs) in genes derived from the WES or WGS data on the selected phenotypes across ancestries (Methods). In total, we identified 1,359 significant (p≤1×10^-8^)^19^ gene-phenotype associations, of which 87.4% (1,188) were significant in both the WES-PheWAS and WGS-PheWAS (representing 184 binary and 1,004 quantitative association signals), 4.9% (66) were significant only in the WES-PheWAS (representing 15 binary and 51 quantitative association signals), and 7.7% (105) were significant only in the WGS-PheWAS (representing 23 binary and 82 quantitative association signals) across the 10 non-synonymous models (Table S13).

There was generally high correlation between the −log_10_(p-values) derived from WES and WGS (Spearman’s rank correlation coefficient = 0.95, p < 2.2×10^-16^) (Fig. S6). Across both binary and quantitative traits, there were 29 genes with significant (p ≤ 1e-8) associations unique to the WGS results and 20 genes with significant (p ≤ 1e-8) associations unique to the WES results (Fig. S7). Three of the genes uniquely associated with either technology were in the MHC region: *VWA7* (WES) and *HLA-C* and *C2* (WGS). In terms of associations, there were 105 (23 binary, 82 quantitative) gene-phenotype pairs significant in WGS but not WES, and 66 (15 binary, 51 quantitative) significant in WES but not WGS (Table S12). We observed that fewer than 3.3% of gene-phenotype pairs had an absolute difference in Phred scores (−10xlog_10_[p-values]) of greater than 5 units and less than 0.56% greater than 10 units (i.e., 1-order of magnitude) (Fig. S8). Across the 14,130,325 gene-phenotype associations (significant and non-significant) there were 54,818 (49,762 binary, 5,056 quantitative) gene-phenotype associations with greater than a 10 unit difference in Phred scores, that achieved a lower p-value in the WGS results, and 23,687 (19,607 binary, 4,080 quantitative) associations that achieved a lower p-value in the WES results (Fig. S9).

We identified 95 significant genotype-phenotype associations with 15 genes recurrently mutated in clonal haematopoiesis and myeloid cancers as described previously^67^, potentially driven by somatic qualifying variants. Of these, 70 were detected by both technologies, 14 were unique to the WES results and 11 were unique to the WGS results. Associations unique to WGS included an association between protein truncating variants in *TET2* and first occurrence phenotype: Source of report of D72 (other disorders of white blood cells) (WGS p-value = 3.62·10^-13,^ binary odds ratio (OR) = 8.08, 95% confidence interval (95% CI) = 5.02 to 12.40; WES p-value = 4.23·10^-7^, OR = 6.18, 95% CI = 3.26 to 10.70). We also found an association between protein truncating and predicted damaging missense variants in *SRSF2* and reticulocyte percentage (WGS p-value = 1.92·10^-6^, beta = 0.30, 95% CI = 0.17 to 0.42; WES p-value = 3.7·10^-18^, beta = 0.60, 95% CI = 0.47 to 0.74) only significant in the WES results (Table S12).

Overall, we found that the association results between the WES and WGS DRAGEN datasets are highly correlated. Nevertheless, we identified examples of genes that contain coding regions/sites where coverage is underrepresented in WES and improved in WGS, resulting in improved association statistics in the WGS versus WES results. One example is *PKHD1* for which we identified associations between rare variant collapsing models and three quantitative phenotypes that were more significant in WGS than WES: gamma-glutamyl transferase (GGT) (WES p-value = 4.63×10^-18^, beta = 0.19, 95% CI = 0.15 to 0.24; WGS p-value = 1.24×10^-19^, beta = 0.20, 95% CI = 0.16 to 0.24), creatinine (WES p-value= 3.85×10^-10^, beta = −0.04, 95% CI = −0.06 to −0.03; WGS p-value = 2.14×10^-12^, beta = −0.05, 95% CI = −0.06 to −0.03) and cystatin C that only achieves significance (p<1×10^-8^) in the WGS data (WES p-value = 3.02×10^-8^, beta = −0.05, 95% CI = −0.07 to −0.03; WGS p-value= 3.04×10^-9^, beta = −0.04, 95% CI = −0.06 to −0.03) (Table S12). Another example is *LPA* where protein-truncating variants are statistically unequivocally associated with lipoprotein A levels in both datasets; however, achieving lower p-values for WGS (WES p-value = 1.92×10^-75^, beta = −0.40, 95% CI = −0.44 to −0.36; WGS p-value = 6.97×10^-86^, beta = −0.40, 95% CI = −0.44 to −0.36) (Table S12). For both *PKHD1* and *LPA,* we observed that the number of samples with ≥10X coverage drops in the WES compared to WGS DRAGEN dataset at specific coding region (CDS) sites/exons (Fig. S10). For example, for *LPA* only ∼39% of samples achieve ≥10X coverage across exons 10-15 (bp: 160,612,932 −160,628,366). These select examples demonstrate settings where the value of WGS improves the ability to ascertain qualifying variants for collapsing analyses and provide a better representation of phenome-wide associations in genes that contain regions with drops in coverage in WES data. From our previous work with the UKB exomes, of the 18,762 studied protein-coding genes we identified 563 genes where only 95% of the protein-coding sequence had on average <10x coverage, 262 of these genes had only 50%, and 133 were as extreme as only 5%^19^. This suggests gaps in discovery potential that could be addressed using the coding regions from the UK Biobank whole-genomes.

#### Rare variant collapsing analysis PheWAS of UTRs yields significant associations

To understand the contributions of rare untranslated region (UTR) variants to human phenotypes, we used the UKB DRAGEN WGS data to compile ∼13.4 million rare variants (minor allele frequency (MAF) < 0.1 %) from both 5’ and 3’ UTRs of protein-coding genes across participants of selected ancestries (EUR, ASJ, AFR, EAS and SAS). We then performed two kinds of multi-ancestry collapsing PheWASs: UTR-only and UTR combined with coding variants.

For the UTR-only collapsing PheWASs, we tested the aggregate effect of UTR qualifying variants on 687 binary and 64 quantitative phenotypes for 5’ UTR only, 3’ UTR only, and 5’ and 3’ UTRs combined. Each of these was run using 6 different collapsing analysis models to capture a range of MAF and CADD^68^ thresholds, and any UTR defined sites that also overlapped with a reported protein-coding site were omitted (Methods).

Using the currently defined collapsing models, we observed a total of 65 significant (P < 1×10^-8^) associations (1 binary and 64 quantitative trait associations) comprising 35 unique genes and 36 unique phenotypes (Figure 6 and Table S13). Of these, 29 (45%) signals were not significant in our WGS-based coding collapsing analyses. One binary signal and 7 quantitative signals were independently significant in both 5’ and 3’ UTR models for those relationships.

**Figure 6:**
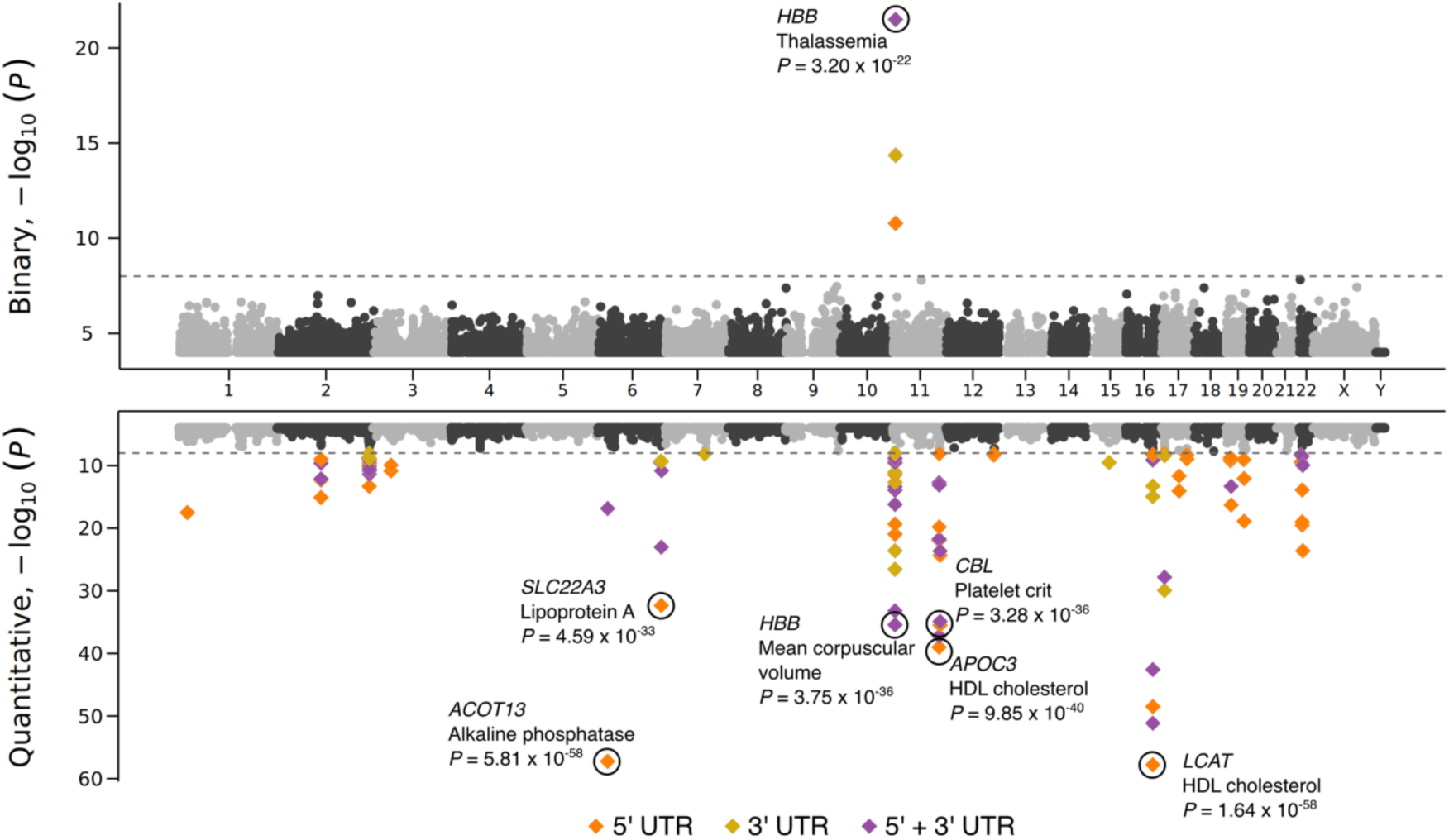
UTR-based collapsing analysis. Miami plot of UTR-based associations for binary (top) and quantitative (bottom) phenotypes across all six collapsing models. 5’, 3’ and 5’ and 3’ combined associations were represented in different colors. The significant associations with P-value < 1 x 10^-30^ were labelled. To understand whether combining UTR and CDS variants helps boost the signals identified by CDS variants only, we performed collapsing analysis using CDS and 5’ and 3’ UTR variants together. For CDS and UTR combined regions, we considered rare PTVs (MAF < 0.1%) from CDS and all variants with MAF < 0.1% from UTR regions. We categorized UTR variants into –two qualifying variant models based on their MAF (i.e., 0.002% and 0.1%- (Method).

For the UTR combined with coding collapsing PheWASs, we tested the combined effect of rare UTR variants and exonic PTVs on the same phenotypes. Each was run using two different collapsing analysis models. We observed 26 and 149 significant (P < 1×10^-8^) associations for binary and quantitative phenotypes respectively (Table S13). We compared these results to our WGS-based coding collapsing PheWAS results. Nine phenotypes that achieved significance in the UTR combined with coding PheWASs, were not significant in the coding-only collapsing PheWAS, suggesting that the coding variant association was boosted by incorporating UTRs (Table S13). For example, rare variants in combined CDS and UTR regions of *NWD1* are significantly associated with kidney calculus (p-value = 9.24·10^-9^, OR = 1.68). The association did not achieve significance (p<1×10-8) in the coding-only collapsing PheWAS or the UTR only collapsing PheWASs, but only when these two regions were combined (Table S14). Our models indicate that the signal is likely to be mostly driven by rare 3’ UTR variants (Table S14) although the 1,220 qualifying variants were distributed throughout *NWD1* CDS and UTR. A recent publication^69^ identified an association between the common synonymous variant rs773852 (MAF = 0.4) and kidney calculus. Our study extends this into rare coding and non-coding variants and demonstrates the value of WGS in identifying non-coding phenotype associations.

#### Phenotypic effects of structural variants

Most of the statistically significant associations that were identified in the previous UKB 150K release^15^ from the WGS consortium have also been identified in the current release. The new UKB 500K release, however, allows the identification of rarer structural variants and assesses their detectable impact on disease and other UK Biobank phenotypes. We present how the full release allows more detailed analysis than the preliminary release, anchoring on genes and variants that have a well-established association with phenotype.

Genes are typically overlapped by several SVs. Previously^15^ we highlighted an association of non-HDL cholesterol with a 14,154 base pair deletion overlapping *PCSK9*, a proprotein convertase which is involved in the degradation of LDL receptors in the liver. In the current release 13 SVs overlapping coding exons in *PCSK9* are found, carried by 163 individuals, bringing the total number of PCSK9 pLOF carriers to 1,124 The previously reported SV is the most common of the 13 variants and is carried by 111 individuals. Carriers of this deletion have markedly (1.22 s.d.) lower levels. Carriers of the other deletions in *PCSK9* similarly had lower levels of non-HDL cholesterol (collectively averaging 0.51 s.d.).

Another example is a 5,200bp deletion on chr12:56,451,164-56,456,364, deleting all four coding exons of *MIP* while largely keeping intact its 5’ UTR region and not affecting other genes. *MIP* encodes the Major intrinsic protein of the lens fiber. Rare deleterious missense and loss-of-function variants in this gene have been associated with autosomal dominant cataract^70,71^ disease; 15 carriers of this deletion are present in the UKB and all belong to the NFE population. We find a strong association with cataracts in our data (OR=25.3, p-value=6.3·10^-7^, MAF = 0.0015%).

The ACMG^72^ recommends reporting actionable genotypes in a list of genes associated with diseases that are highly penetrant and for which a well-established intervention is available. We previously reported^15^ that 4.1% of individuals in the UKB carry an actionable SNP or indel genotype. We find that an additional 0.60% of individuals carry an SV that is annotated to cause a loss of function in a gene annotated to be autosomal dominant LoF/P/LP. While further validation of these markers^73^ is needed, this represents a 14.8% increase in the number of individuals with an actionable genotype were all these variants found to be LoF/P/LP.

ClinVar^74^ is an archive for interpretations of clinical significance of variants for reported conditions. The latest release of ClinVar has 2,256,088 records, of which only 4,062 are SVs; 458 SVs presented here matched 486 (12.0%) of the 4,062 SVs in ClinVar. As expected, variants annotated as benign or likely benign are generally carried by more individuals in the general population setting than those that are annotated as pathogenic or likely pathogenic (Table S15). The scale of this population sample cohort coupled with access to rich medical history allows us to assess the likely clinical impact of many of these variants across a number of phenotypes and thus reassess the ClinVar classification.

Most SVs annotated as pathogenic in ClinVar and found in our data are very rare (MAF< 0.01%,Table S15). One example of a ClinVar variant is a 52 base pair deletion on chr19:12,943,750-12,943,802 in the first exon of *CALR* gene resulting in a stop gain. The variant is a recurrent somatic mutation^75–77^ and annotated in ClinVar as pathogenic for primary myelofibrosis and thrombocytoma carried by 47 individuals in the NFE population and one individual in the AFR population. We find that the variant associates with measures of platelet distribution; most strongly with platelet width, effect 2.02 s.d. (95% CI 1.72-2.34, p-value=3.1×10^-38^). Interestingly this variant is also found in our SNP/indel call set, but is not found in the WES data, despite being exonic.

Although most ClinVar variants are very rare within the UK Biobank some of the variants have higher frequency in the sub-cohorts, an example of this is a 2,502 bp deletion on chr2:151,645,755-151,648,057 that deletes exon 55 of *NEB*. This deletion has been associated with nemaline myopathy, is known to have arisen from a single founder mutation^78^ and is carried by 33 individuals in the cohort; 17 of which belong to the ASJ sub-cohort. A second example is a 613bp deletion on chr11:5,225,255-5,225,868 that deletes the first of three exons of *HBB*. The deletion is carried by 19 individuals all belonging to the SAS cohort. The deletion has been annotated in ClinVar to be clinically significant for beta Thalassemia and we find it to associate with a 1.96 s.d. (95% CI 1.49-2.43, p-value=5.4·10^-16^) decrease in Haemoglobin concentration.

## Discussion

The UKB WGS project offers a groundbreaking opportunity to explore human genetic variation and its application to disease research. The vast dataset generated in this study will advance our understanding of human genetics and significantly impact drug discovery and development, disease risk assessment and precision medicine applications on a global scale. Furthermore, this work will provide essential insights to help understand the contribution of rare non-coding variation to human biological variation and will facilitate the translation of human genetics into therapies over the next decade. This dataset will also enable advances in evolutionary biology such as understanding of *de novo* variation, evaluation of polygenic adaptation and improved inference from ancient DNA. Moreover, lessons learned during this endeavor, by successfully addressing scientific, technical, and organizational challenges by leveraging public-private partnerships, provide a blueprint and benchmark for future population-scale studies.

UKB WGS was crucial to identify an 18.8-fold increase of variants over the imputed array and over 40-fold increase to WES, providing an expanded panoramic view of genomic landscape. This is consistent with multiple studies that highlight the power of WGS versus WES for identifying coding variants^5^, especially considering the decreased cost of WGS over time^6^. In accordance with previous efforts^15,16^ this information can be used to identify regions that have a lower tolerance to variation. From Figure 4, WGS clearly allowed us to identify more genes harboring pLoF/P/LP variants in more carriers, which offers more opportunities of evaluating gene targets in LoF heterozygous carriers or even human “KO”. WGS also allowed us to find a large number of clinically relevant and disease-associated structural variants. The incremental benefit of variant identification from WGS has implications on the design and budgeting of future population size studies, especially in identifying variants largely over-represented in specific non-NFE groups.

From cross-ancestry meta-analysis, we confirmed known associations and identified novel ones with new indications and/or in non-European ancestries. Even though non-NFE ancestries had smaller sample sizes, 82 meta-GWS associations were found significant only in non-NFE ancestries (Table S9), likely driven by selection pressure from regional-specific environment factors. For example, *HBB* and *G6PD*’s missense causal variants for SCD and anaemia were >1500x more common in AFR vs. NFE, due to their protection against severe malaria and the fact that 95% of malaria cases occur in African^80^. In contrast, a Thalassemia-causing *HBB* LoF mutation (rs11549407-A) and splice site variant (rs33915217-G) were most prevalent in NFE and SAS. These variants are rare in AFR and have no reported protective effects against malaria or other infectious diseases endemic to Africa. While a *HBB* nonsense variant was detected in WES (AF=0.003%) but more enriched in WGS (AF=0.005%), the splice site variant was exclusively detected in WGS (not in WES nor in imputed array genotypes), again highlighting WGS’ unique value.

There is consensus among the pharmaceutical industry that human genomic evidence increases the FDA approval rate at least 2-fold^81–84^. But current human genomic reference/biobank data do not reflect the diversity of human populations i.e., are still dominated by European ancestries^85^ thus limiting the detection of variation specific to non-European regions and leading to a fundamental bias in the understanding of the genetic basis of disease in diverse populations. In recent years, there has been increasing awareness of the inequities in the representation of global populations from non-European descent in clinical trials and basic clinical research. Attempts to address these inequities will reduce health disparities that affect individuals of non-European or admixed ancestries. There is a strong need for more diverse and deeper human multiomic data, which will generate novel insights as a complement to other pre-clinical models for drug discovery and development^86^.

To understand the impact of rare variants captured by WGS on human disease we present a series of examples using collapsing analysis including non-coding variants and coding variants that are not well covered by standard WES. Our observation that WGS can boost significance for certain genetic associations compared to WES in a collapsing analysis PheWAS context is consistent with other studies that show better coverage (and therefore better sensitivity to call variants) in WGS compared to WES for particular genes^79^. Genes that have improved coverage in WGS include *LPA*, which has been suggested as a gene that could be screened for cardiovascular risk prediction^87^, demonstrating potential clinical utility of WGS. While we restricted collapsing analyses to samples who had both WES and WGS available to allow a closer comparison of the technologies, nevertheless some technical differences remain (e.g., differences in read length and average coverage depth of coding regions) that could contribute to the difference in signals. Defining qualifying variants in non-protein-coding regions remains challenging. *In silico* predictions of variant functional effect are less accurate in non-protein-coding regions than protein coding regions. Additionally, biological effects of variation in non-protein-coding regions are likely to be on average more modest than those in protein-coding regions. Therefore, in this study augmenting collapsing analysis signals in coding regions with UTRs dilutes the signals for many phenotypes. Nevertheless, our observation of significant associations in UTRs, and a few phenotypes for which adding UTR boosts coding signals, demonstrates the great potential of using this dataset to explore disease-relevant rare variant associations in neglected non-coding regions. Next steps could include further refining the non-coding qualifying variant definitions with additional filters, expanding to other phenotypes, and other classes of non-coding regions. In the UK Biobank, additional data modalities provide a valuable opportunity to discriminate functionally important variants and therefore refine qualifying variant criteria. For example, a recent study using Olink plasma proteomics data in the UK Biobank boosts signals by combining protein quantitative trait loci with protein-truncating variants in collapsing analyses^88^.

The study presented here is the largest WGS-based genetic study performed to date. We have provided examples where we show that combining WGS data with the rich phenotypic data in the UK Biobank gives new insights into the complex relationship between human variation and sequence variation. We believe that further analysis of the data will be invaluable. This resource can not only facilitate improved imputation performance for rare variants in individuals across five different ancestries ^14,15^, but may also be useful for describing variation in complex regions, such as HLA, KIR, and red blood cell antigen systems, and serve as a gold standard for future population scale studies. We are confident that leveraging the combined expertise of scientists worldwide will lead to new insights that will meaningfully impact our understanding of human disease biology and thereby advance the search for safe and effective medicines.

## Data availability

WGS data can be accessed via the UK Biobank research analysis platform (RAP): https://ukbiobank.dnanexus.com/landing. The Research Analysis Platform is open to researchers who are listed as collaborators on UKB-approved access applications. Allele frequency browser is available at https://afb.ukbiobank.ac.uk/. Rare variant collapsing analysis association statistics are available through the AstraZeneca Centre for Genomics Research (CGR) PheWAS Portal (http://azphewas.com/). SV association data is available at https://www.decode.com/summarydata/. Human reference genome GRCh38, http://ftp.1000genomes.ebi.ac.uk/vol1/ftp/technical/reference/GRCh38_reference_genome/. GIAB WGS samples https://ftp-trace.ncbi.nlm.nih.gov/ReferenceSamples/giab/data/ ENSEMBL https://m.ensembl.org/info/data/mysql.html.

## Acknowledgements

This research has been conducted using the UK Biobank Resource (applica}on no. 52293). This research has been conducted with the support of Wellcome (grant nos. 218880/Z/19/Z to Amgen, 218723/Z/19/Z to AstraZeneca, 218789/Z/19/Z to GSK, 219414/Z/19/Z to Janssen and 219603/Z/19/Z to UK Biobank), UK Research and Innova}on (Innovate UK grant nos. 105637 to Amgen, 105635 to AstraZeneca, 105634 to GSK and 105636 to Janssen), Medical Research Council award for Vanguard programme (grant no. MC_PC_17223). We would like to acknowledge the contribu}ons of the laboratory and wider support teams at UK Biobank and thank all the 500,000 par}cipants of the UK Biobank. We thank Zhuoyi Huang, Shyamal Mehtalia, Rami Mehio, and the Illumina DRAGEN team for their technical support with DRAGEN data processing and aggregate calling. We thank James Jacob from Amazon Web Services EMEA SARL for the design of the DRAGEN data processing pipeline. We thank Fiona Middleton, Rob Wenier, and Brian Smalley from AstraZeneca for their technical leadership.

