## Supplemental Note for "Whole-genome sequencing of half-a-million UK Biobank participants"

#### Authorship

##### AstraZeneca

Keren Carss[1], Eleanor Wheeler[1], Kousik Kundu[1], Fengyuan Hu[1], Quanli Wang[2], Ryan Dhindsa[2], Sri V. V. Deevi[1], Kieren Lythgow[1], Peter H. Maccallum[1,3], Karyn Mégy[1], Jonathan Mitchell[1], Sean O'Dell[1], Amanda O'Neill[1], Katherine R. Smith[1], Haeyam Taiy[1], Menelas Pangalos[4], Ruth March[5], Sebastian Wasilewski[1], Slavé Petrovski[1,6,7]

1)Centre for Genomics Research, Discovery Sciences, BioPharmaceuticals R&D, AstraZeneca, Cambridge, UK
2)Centre for Genomics Research, Discovery Sciences, BioPharmaceuticals R&D, AstraZeneca, Waltham, MA, USA
3) ELIXIR, Wellcome Genome Campus, Hinxton, Cambridge, CB10 1SD, UK
4) BioPharmaceuticals R&D, AstraZeneca, Cambridge, UK
5) Precision Medicine & Biosamples, Oncology R&D, AstraZeneca, Cambridge, UK.
6) Department of Medicine, University of Melbourne, Austin Health, Melbourne, Victoria, Australia
7) Epilepsy Research Centre, University of Melbourne, Austin Health, Melbourne, Victoria, Australia.

Author contributions:
Study conceptualization, project coordination and consortium leadership: MP, RM, SW, and SP. Design and implementation of statistical analyses and data interpretation: KC, EW, KK, FH, QW, RD, KRS, JM, and SP. Data processing: FH, QW, SD, KL, PM, KM, SO, AO, and HT. Figure preparation: EW and KK. Manuscript writing: KC, EW, KK, RD, and SP. All the authors reviewed the manuscript.

##### deCODE/Amgen

Bjarni V. Halldorsson[1,2], Hannes P. Eggertsson[1], Kristjan H.S. Moore[1], Hannes Hauswedell[1], Ogmundur Eiriksson[1], Gunnar Palsson[1], Marteinn T. Hardarson[1,2], Asmundur Oddsson[1], Brynjar O. Jensson[1], Snaedis Kristmundsdottir[1,2], Vinicius Tragante[1], Arnaldur Gylfason[1], Pall I. Olason[1], Margret Asgeirsdottir[1], Sverrir T. Sverrisson[1], Brynjar Sigurdsson[1], Sigurjon A.

Gudjonsson[1], Gunnar T. Sigurdsson[1], Gardar Sveinbjornsson[1], Droplaug N. Magnusdottir[1], Steinunn Snorradottir[1], Kari Kristinsson[1], Emilia Sobech[1], Gudmar Thorleifsson[1], Frosti Jonsson[1], Pall Melsted[1,3], Ingileif Jonsdottir[1,5], Thorunn Rafnar[1], Hilma Holm[1], Hreinn Stefansson[1] Jona Saemundsdottir[1], Daniel F. Gudbjartsson[1,3], Olafur T. Magnusson[1], Gisli Masson[1], Unnur Thorsteinsdottir[1,5], Agnar Helgason[1,6], Hakon Jonsson[1], Patrick Sulem[1], Kari Stefansson[1]

1 deCODE genetics / Amgen Inc., Sturlugata 8, Reykjavik, Iceland
2 School of Technology, Reykjavik University, Reykjavik, Iceland
3 School of Engineering and Natural Sciences, University of Iceland, Reykjavik, Iceland
4 Landspitali-University Hospital, Reykjavik, Iceland
5 Faculty of Medicine, School of Health Sciences, University of Iceland, Reykjavik, Iceland
6 Department of Anthropology, University of Iceland, Reykjavik, Iceland

Author contributions:
Study conceptualization, project coordination and consortium leadership: BVH, FJ, OTM, DFG, GM, UT, AH, PS and KS DNA sequencing and sample preperation DNM, SS, KK, ES- JS ,OTM. Design and implementation of statistical analyses and data interpretation: BVH, HPE, KHSM, OE, GP, AO, BOJ, VT, GSD, GT, PM, IJ, TR, HHo, HS, HJ Data processing: BVH, HPE, HHa, SK, AG, PIO, MA, STS, BS, SAG, GTS, GM Figure preparation: KHSM and MTH. Manuscript writing: BVH and KS. All the authors reviewed the manuscript.

#### GSK

Jimmy Liu[1], Yancy Lo[1], Jatin Sandhuria[2], Tom G. Richardson[3], Laurence Howe[3], Chloe Robins[1], Dongjing Liu[1], Patrick Albers[3], Mariana Pereira[3],Daniel Seaton[3], Yury Aulchenko[3], John Whittaker[3], Manolis Dermitzakis[3], Toby Johnson[3], Jonathan Davitte[1], Erik Ingelsson[4], Robert Scott[3], Adrian Cortes[3]

Affiliations:
1. Human Genetics, Genomic Sciences, GSK, Collegeville, PA, USA
2. R&D Data Science & Data Engineering, Collegeville, PA, USA
3. Human Genetics, Genomic Sciences, GSK, Stevenage, UK

Author contributions
Study conceptualization, project coordination and consortium leadership: JW, RS and AC.
Design and implementation of statistical analyses and data interpretation: JL, YL and AC.
Data processing: JL, YL, TGR, JD and AC.
Figure preparation: JL, YL and AC.
Manuscript writing: JL, YL, RS and AC.
All the authors reviewed the manuscript.

#### Johnson & Johnson

Liping Hou[1], Julio Molineros[1], Yanfei Zhang[1], Alexander H Li[1], Evan H Baugh[1], Elisabeth Mlynarski[1], Abolfazl Doostparast Torshizi[1], Gamal Abdel-Azim[1], Brian Mautz[1], Karen Y. He[1], Xingjun Liu[1], Antonio R. Parrado[1], Dongnhu Truong[1], Mohamed-Ramzi Temanni[1], Christopher D. Whelan[2], Letizia Goretti[3,7], Najat Khan[4], Tommaso Mansi[1], Guna Rajagopal [5,8], Mary Helen Black[1,9], Trevor Howe[6] & Shuwei Li[1]

1 AI/ML, R&D Data Science & Digital Health, Johnson & Johnson Innovative Medicine, PA US
2 DS NS, R&D Data Science & Digital Health, Johnson & Johnson Innovative Medicine, MA US
3 External Innovation, R&D Discovery, Product Development & Supply, Johnson & Johnson Innovative Medicine
4 R&D Data Science & Digital Health, Johnson & Johnson Innovative Medicine, PA US
5 Computational Science, R&D Discovery, Product Development & Supply, Johnson & Johnson Innovative Medicine
6 External Innovation, R&D Data Science & Digital Health, Johnson & Johnson Innovative Medicine, PA US
7 current address: Alia Therapeutics SRL, Via Vincenzo Gioberti 8, 20123 Milano
8 current address: Samsara BioCapital, 628 Middlefield Road, Palo Alto, CA 94301
9 current address: Foresite Labs, One Boston Place Suite 4010. Boston, MA 02108

Author contributions:
Study conceptualization, project coordination and consortium leadership: TH, SL, MHB, GR, LG, TM and NK. Design and implementation of statistical analyses and data interpretation: SL, LH and JM. Data processing: LH, JM, YZ, AL, EB, EM, AT, GA, BM, KH, XL, AP, DT, MT. Figure/Table preparation: LH, JM, YZ and SL. Manuscript writing: SL, TH and MHB. All the authors critically reviewed the manuscript.

#### Sanger (Velsera Seven Bridges)

Shaheen Akhtar[1], Siobhan Austin-Guest[1], Robert Barber[1], Daniel Barrett[1], Tristram Bellerby[1], Adrian Clarke[1], Richard Clark[1], Maria Coppola[1], Linda Cornwell[1], Abby Crackett[1], Joseph Dawson[1], Callum Day[1], Alexander Dove[1], Jillian Durham[1], Robert Fairweather[1], Marcella Ferrero[1], Michael Fenton[1], Howerd Fordham[1], Audrey Fraser[1], Paul Heath[1], Emily Heron[1], Gary Hornett[1], Lena Hughes-Hallett[1], David K Jackson[1], Alexander Jakubowski Smith[1], Adam Laverack[1], Katharine Law[1], Steven R Leonard[1], Kevin Lewis[1], Jennifer Liddle[1], Alice Lindsell[1], Sally Linsdell[1], Jamie Lovell[1], James Mack[1,] Henry Mallalieu[1], Irfaan Mamun[1], Neil Marriott[1], Ana Monteiro[1], Leanne Morrow[1], Barbora Pardubska[1], Alexandru Popov[1], Carol E Scott[1], Lisa Sloper[1], Jan Squares[1], Ian Still[1], Oprah Taylor[1], Sam Taylor[1], Jaime M Tovar Corona[1], Elliott Trigg[1], Valerie Vancollie[1], Paul Voak[1], Danni Weldon[1], Alan Wells[1], Eloise Wells[1], Mia Williams[1], Sean Wright[1], Ahmet Sinan Yavuz[2], Jelena Randjelovic[2], Nevena Miletic[2], Lea Lenhardt Ackovic[2], Marijeta Slavkovic-Ilic[2],Mladen Lazarevic[2] Diana Rajan[1], Louise Aigrain[1], Nicholas Redshaw[1], Michael Quail[1], Lesley Shirley[1], Scott Thurston[1],Peter Ellis[1], Laura Grout[1], Natalie Smerdon[1], Emma Gray[1], Richard Rance[1], Cordelia Langford[1], Ian Johnston[1]
We would like to acknowledge the contributions of the wider support teams at Sanger.

We would like to acknowledge the contributions of the bioinformatics, engineering, product, scientific engagement and support teams at Velsera, formerly Seven Bridges.
Study conceptualization, project coordination and consortium leadership: CL and IJ
DNA sequencing and sample preparation: SA, SAG, RB, DB, TB, AC, RC, MC, LC, AC, JD, CD, AD, JD, RF, MF, MF, HF, AF, PH, GH, LHH, AJS, AL, KL, AL, SL, JL, HM, JM, IM, NM, AM, LM, BP, AP, LS, JS, IS, OT, ST, ET, VV, PV, DW, AW, EW, MW, SW, LG, NS, EG, RR, CL and IJ
Data processing: DKR, CES, JMTC, JD, KL, SRL, JL, ASY, JR, NM, LLA, MS-I and ML
Methods writing: IJ, DR, DKR, CES, ASY, NM and JR
Affiliations:

1. Wellcome Sanger Institue, Wellcome Genome Campus, Hinxton, Cambridge, CB10 1SD, UK
2. Velsera, Charlestown, MA, US

#### UK Biobank

Rory Collins, Mark Effingham, Naomi Allen, Jonathan Sellors, Ben Lacey, Simon Sheard, Mahesh Pancholi, Caroline Clark, Lucy Burkitt-Gray, Samantha Welsh, Daniel Fry, Rachel Watson, Lauren Carson, Alan Young

Study conceptualization, project coordination and consortium leadership: RC, ME, NA, JS, SS, CC, DF, LC, LBG
Sample preparation: SW, DF, RW
Data processing: CC, LBG, AY, MP
Manuscript writing: BL

### Methods

#### Ethics statement

The UKB phenotype and genotype data were collected following an informed consent obtained from all participants. The North West Research Ethics Committee reviewed and approved UKB's scientific protocol and operational procedures (REC Reference Number: 06/MRE08/65). Data for this study was obtained and research conducted under the UKB applications license numbers 24898 and 68574.

#### Sequencing data set

Sequencing was made possible by a public-private partnership between UK Biobank, UK Research and Innovation, Wellcome and four industry partners (Amgen, AstraZeneca, GSK and Johnson & Johnson). From the total UK Biobank cohort of 503,310 participants, 807 had withdrawn consent prior to the start of this study and 10,949 had no suitable sample for sequencing. Sequencing was performed in two centers (deCODE facility in Reykjavik, Iceland and the Wellcome Sanger Institute (Sanger) in Cambridge, UK). 50,010 samples were sequenced as part of the Vanguard phase of this project. Samples from an additional 441,544 individuals were prepared for sequencing. In total 492,729 samples from 491,554 individuals were sequenced as part of either this study or its Vanguard phase. The sequence data included 1,175 replicates, where 185 were included as accidental replicates and 990 as technical replicates. The sequencing of 914 participants failed due to either insufficient or poor-quality DNA, for a total of 490,640 successfully sequenced individuals. An additional 91 individuals withdrew consent from the time of start of sequencing until commencement of joint calling. The remaining 490,549 successfully sequenced primary samples 20 of the accidental replicates and the 990 technical replicates were used for joint calling with GraphTyper[1], for a total of 491,559 samples.
Out of the 490,549 primary samples, 49,934 samples were sequenced as part of the Vanguard phase, 193,093 and 247,522 samples were sequenced by Sanger and deCODE, respectively.

#### Sequence data processing

Three commensurate bioinformatics pipelines were developed (Supplementary Note 3: Sequence processing pipeline). All pipelines were designed to comply with the principles of functional equivalence[2] and the Broad Institute Best Practices workflows[3], which were used on different sets of samples.
The key components of the per sample data product provided by these pipelines were a bwa mem[4], GRCh38[5] (with alt contigs plus additional decoy contigs and HLA genes) reference, aligned cram file (containing original instrument basecall quality values), a crai index file, a GATK gVCF file and GATK BQSR quality recalibration table.

#### SNP and indel calling with GraphTyper

In addition to the 491,559 sequenced samples from the UKB, 7 samples from the Genome In A Bottle consortium[6] were included as quality controls. Metrics of call set accuracy are shown in Table S4.
Prior to running GraphTyper[1] we preprocessed all input CRAI indices by extracting a large single file containing all CRAI index entries with sample_id for a 20kb window (with 1 kb padding at each side of the region) for all samples. For each region, we then created a

chopped CRAI for each sample by processing the large file for the corresponding region, substantially reducing the amount of CRAI index entries read.
Further, we created a sequence cache of the reference FASTA file using the `seq_cache_populate.pl` script distributed with samtools[2] 1.9. In each region we copied the corresponding sequence cache to the local disk and used it for reading the CRAM files by setting the `REF_CACHE` environment variable.
We ran GraphTyper[1] (v2.7.5) using the `genotype` subcommand. The full command we ran was in the format:

```
graphtyper genotype ${UKBIO_REFERENCE}
 --sams=${SAMS}
 --sams_index=${CRAI_TMP}/crai_filelist.txt
 --avg_cov_by_readlen=${COVERAGES}
 --region=${REGION}
 --threads=${THREADS}
 --verbose
```

Where `UKBIO_REFERENCE` is the `GRCh38_full_analysis_set_plus_decoy_hla` FASTA sequence file, `SAMS` is a list of all input BAM/CRAM files, `CRAI_TMP` is a path to the chopped CRAI files on the local disk, `COVERAGES` is the coverage divided by the read length for each input file, `REGION` is the genotyping region and `THREADS` is the number of threads to use.

##### Running time

All jobs were run using 12 cores with 66GB of reserved RAM. Approximately 2% of jobs had to be rerun using either 48 cores or 132GB of reserved RAM. A few jobs required up to 48 cores and 350GB of RAM. Total reserved CPU time on cluster, including reruns, was 37.4M CPU hours and total effective compute time was 31.0M CPU hours. The difference in these numbers is explained by the fact that not all cores reserved for the program could be utilized simultaneously at all times.

#### SV calling with Manta and GraphTyper

We ran a structural variant (SV) genotyping pipeline similar to the one we had previously applied to 49,962 Icelanders[7] and in for the first release of the WGS dataset[8], with the modification that the DRAGEN SV caller was run instead of Manta. In summary, we ran the SV caller within DRAGEN v3.7.8 to discover SVs on all 490,241 individuals in the genotyping set. We used svimmer[7] to merge these different SV datasets and we called the resulting SVs using GraphTyper[1,7] version 2.7.5.
A total of 2,758,898 variants were called of which variants were 1,926,132 annotated as PASS. For each variant 4 different models for genotyping were run, a variant was considered reliable if one of them annotated the variants as PASS. When multiple models PASSed, we selected the model with the lowest duplicate error rate among them. Variant counts are presented for variants annotated by GraphTyper as PASS, unless otherwise noted.

#### Sequence data processing and single sample variant calling with DRAGEN

UK Biobank whole genome sequencing data were processed at AstraZeneca as previously described[9]. Illumina DRAGEN Bio-IT Platform Germline Pipeline v3.7.8 was run within Amazon

Web Services cloud platform. The sequence reads were aligned to the GRCh38 graph genome reference and SNVs and small indels were called at a sample level.

The full command we ran was in the format:

```
/opt/edico/bin/dragen \
--bam-input ${SAMPLE_ID}.bam \
--cnv-enable-self-normalization true \
--enable-cnv true \
--enable-cyp2d6 true \
--enable-duplicate-marking true \
--enable-map-align true \
--enable-map-align-output true \
--enable-sort true \
--enable-sv true \
--enable-variant-caller true \
--intermediate-results-dir /scratch/${OUT_DIR} \
--output-directory /${OUT_DIR} \
--output-file-prefix ${SAMPLE_ID} \
--output-format CRAM \
--qc-coverage-count-soft-clipped-bases true \
--qc-coverage-ignore-overlaps true \
--qc-coverage-region-1 wgs_coverage_regions.hg38_minus_N.interval_list.bed \
--qc-coverage-region-2 acmg59_allofus_19dec2019.GRC38.wGenes.NEW.bed \
--qc-coverage-region-3 CGR_adjusted_CCDS_r22_merged.bed \
--qc-coverage-reports-1 cov_report \
--qc-coverage-reports-2 cov_report \
--qc-coverage-reports-3 full_res \
--qc-cross-cont-vcf
/opt/edico/config/sample_cross_contamination_resource_hg38.vcf.gz \
--read-trimmers polyg \
--ref-dir /reference/hg38_alt_aware \
--repeat-genotype-enable true \
--repeat-genotype-specs variant_catalog.json \
--soft-read-trimmers none \
--vc-emit-ref-confidence GVCF \
--vc-enable-joint-detection true \
--vc-enable-vcf-output true \
--vc-frd-max-effective-depth 40 \
--vc-hard-filter DRAGENHardQUAL:all:QUAL<5.0;LowDepth:all:DP<=1
```

where all the relevant BED files and reference files are available for download from the Illumina webpages (https://developer.illumina.com/dragen/dragen-popgen)

Regarding running time, DRAGEN was run on AWS f1.4xlarge instances (16 cores with 244GB RAM each) equipped with an FPGA accelerator. A few jobs required to be run on f1.8xlarge instances due to memory requirements. The workload took 17.0M CPU hours to compute and another 2M CPU hours to support network transfers between UKB research analysis platform (RAP) and the AWS compute environment. The total volume of data returned to UK Biobank was 12.3 PB.

UK Biobank whole exome sequencing data were processed at AstraZeneca as previously described[10]. Briefly, genomic DNA underwent paired-end 75-bp whole-exome sequencing at Regeneron Pharmaceuticals to an average coverage of 58x using the IDT xGen v1 capture kit and the NovaSeq6000 platform. Illumina DRAGEN Bio-IT Platform Germline Pipeline v3.0.7 was used to align the reads to the GRCh38 genome reference and to call small indels and SNVs (single sample calling).

For both genomes and exomes, small variants were annotated using SnpEff[11] v4.3, Ensembl[12] Build 38.92, REVEL[13], MTR[14], and CADD[15] v1.4.

To define high-quality variants, we applied a number of stringent variant-level quality control (QC) steps as described previously[16] . In brief, the variant-level QC criteria included coverage depth (minimum coverage 10X), genotype and mapping quality scores, DRAGEN variant status, read position rank sum score (RPRS), mapping quality rank sum score (MQRS), alternate allele read proportion for heterozygous calls, proportion of samples failing any of these QC criteria, and gnomAD-related filters.

#### Aggregate variant calling with DRAGEN

DRAGEN Machine Learning Recalibration (MLR) was run on each single sample called with DRAGEN v3.7.8, to recalibrate variant quality and genotype quality with features collected from DRAGEN v3.7.8 alignment and variant calling. To ensure high precision and sensitivity, we assess the sample level accuracy of DRAGEN variant calling using GIAB samples (Table SX5), and use ML recalibrated QUAL=3.0 as the quality cutoff for the aggregated dataset. High confidence regions for GIAB samples are as defined by NIST v4.2.1.

DRAGEN Iterative gVCF Genotyper is utilized to aggregate samples per batch of 1000 and perform genotyping across 490,541 individuals. This dataset covers the autosomes (chromosome 1-22), sex chromosomes (chrX, chrY), mitochondria (chrM) and 3341 ALT contigs of hg38.

In total, we launch 874,000 analyses on Illumina Analytics (ICA) Platform using non-FPGA software instance (16 vCPU 128GB ram for MLR pipeline and 36 vCPU 72 GB for IGG pipeline). The total amount of compute for 500K WGS aggregated dataset is 7.3 million CPU hours, effectively done on ICA in only 72 days.

#### Comparison of SVs to ClinVar

A vcf file containing ClinVar version 20231007 was downloaded.  All variants with an allele in the ref or alt field that had length at least 50bp were considered SVs, resulting in 4,062 SVs. A start position for the SV is given in the vcf file and an end position was computed from the length of the alt allele.  A variant in the SV dataset presented here was considered to match an SV in ClinVar if start and end positions were both within 10bp of each other.

#### Cohort definitions

Individuals were assigned to one of 9 ancestry groups using a random forest classifier trained in the gnomAD[17] v3.1 dataset. Variant loadings for 76,399 ancestry-informative variants from gnomAD were used to project the first 16 principal components onto all UKB WGS samples. A random forest classifier trained on nine known ancestry groups within gnomAD (based on HGDP and 1000 Genomes samples) was then used to calculate ancestry probabilities in the UKB WGS samples. We assigned ancestry labels based on a minimum probability of 0.9, and remaining individuals were assigned as “other”. Population cohorts with over 1,000 individuals were used for genome-wide association analysis.

#### Phenotype data

Phenotype data was ascertained from the UK Biobank Data Showcase. For disease traits we used the first occurrence data (UK Biobank Showcase Category 1712) and analysed 764 ICD-10 codes. For quantitative traits we analysed 64 molecular phenotypes including all blood and urine biochemistry and cell count data (UK Biobank Showcase Category 17518 and 100081) and 7 anthropomorphic traits from the baseline assessment data (UK Biobank Showcase Category 100010). All quantitative traits were rank-based inverse-normal transformed prior to analysis.

#### Single variant association analysis

##### Genotype filtering

Genetic datasets were prepared consistently for each population cohort. The joint VCF files from GraphTyper were converted to biallelic BGEN14 1.2 format files. Variants were excluded based on GraphTyper metrics (AAScore < 0.15, Pass ratio < 0.05, ABhet < 0.175, ABhom < 0.9, QD > 6 and QUAL < 10) as well as per-population cohort metrics (minor allele count < 25, Hardy-Weinberg equilibrium test P < 1e-100, missingness rate > 0.1).

##### Association analysis

Association analysis for SNPs and small indels in all autosomal and chromosome X were performed using REGENIE[18]. For Step 1 of REGENIE, we selected a set of common LD-pruned variants for each population cohort using PLINK (options: --maf 0.01 --indep-pairwise 1000kb 0.1). The total number of variants for Step 1 ranged from 266,859 for the ASJ cohort to 709,479 for the AFR (African) cohort. The resulting predictors were including as covariates in the association analysis of Step 2 of REGENIE, in addition to genotype-derived sex, age at baseline, sequencing centre, and the first 20 genotype ancestry principal components.

We applied a distance-based approach to define a list of associated loci ("top hits") for each phenotype: For each chromosome, if there are variants with $P < 5{\cdot}10^{-8}$, we recursively select the variant with the smallest P-value within a +/-500KB window until there are no remaining variants with $P < 5 \cdot 10^{-8}$. From this list of selected variants, we merge those that are within 1MB of each other into a single locus and select the variant with the smallest P-value as the top hit for that locus. Variants that do not need merging are considered top hits on their own.

To estimate the gain in the number of top hits in the WGS GWAS compared to imputed array GWAS, we calculate the number of top hits for each phenotype using 1) all variants from the WGS GWAS and 2) the subset of variants in the WGS GWAS that were previously genotyped and well-imputed (INFO > 0.3) from the V3 imputed array data. A top hit is considered novel in WGS if it is found in 1) and does not overlap with any top hit in 2). Here, an overlap is defined as the WGS GWAS top hit being within at least a +/-500KB window (or wider if multiple significant variants were merged during the distance-based top hit procedure) of the imputed array GWAS top hit.

##### Putative LoF (Loss-of-function), Pathogenic/Likely pathogenic (P/LP) variant annotation

We identified putative LoF (pLoF) variants in the UKB WGS data and compared with the pLoF variants detected from WES. The pLoF variants were defined as alterations with high function impact (stop lost/gained, start lost, frameshift, splice donor/acceptor) using VEP[19] (release 101, hg38), with gnomAD allele frequency <1%. LOFTEE was used to distinguish high-confidence (HC) pLoF variants from potential annotation artifacts by applying stringent filtering criteria (eg. removing variants predicted to escape nonsense-mediated decay). Only the LOFTEE-predicted HC pLoF variants in the canonical transcript were considered for summary. We also included the ClinVar classified pathogenic and likely pathogenic with assertion criteria and without conflicting classification into the summary of pLoF/P/LP variants and carriers.

#### Region-based PheWAS methods

##### Sample selection

We included individuals from the five ancestry groups with both WES and WGS data available. We applied additional exclusions as previously described[10], excluding samples with VerifyBAMID freemix (a measure of DNA contamination) of more than 4%, where <94.5% of the consensus coding sequence (CCDS release 22) achieved a minimum of 10-fold read depth and where there was a mismatch between self-reported and genetic sex (X:Y CCDS coverage ratios). After QC, there were 460,552 samples for analysis: NFE (N=437,812), ASJ (N=2,671), AFR (N=8,701), EAS (N=2,150), SAS (N=9,218)

##### Phenotypes

We analysed 687 binary First Occurrence phenotypes from the UK Biobank 2022-06 release that had at least 100 cases in UK Biobank and were not among a small number of potentially sensitive phenotypes. We included 64 quantitative phenotypes: blood biochemistry (N=30), blood cell counts (N=28) and physical measures (N=6) height, BMI, systolic blood pressure, diastolic blood pressure, waist circumference, hip circumference. Quantitative phenotypes were inverse-normal transformed before analysis, and phenotypes with less than 20 different values across the included individuals were excluded. Details of binary and quantitative phenotypes studied are provided in Table S10.

##### Models

We performed our previously described gene-level collapsing analysis framework[10]. Briefly, we define high quality qualifying variants (QVs) to create 10 nonsynonymous collapsing models, including 9 dominant models and 1 recessive model, plus an additional synonymous variant model as an empirical negative control (Table S11). We identified QV carriers within each of the 5 ancestry groups across each of the models and compared carriers to non-carriers using the DRAGEN datasets.

For binary traits, we used Fisher's exact two-sided test to compare the difference in the proportion of cases and controls carrying QVs in each gene (Ensembl[12] CCDS public release 22). For quantitative traits we tested the difference in the mean of the phenotype by fitting a linear regression model, correcting for age, sex, 4 genetic principal components and sequencing batch (WES) or sequencing site (WGS). For the dominant collapsing models, we identified carriers of at least one QV in a gene and compared to the noncarriers. For the recessive model, individuals with two copies of QVs in either homozygous or putatively compound heterozygous form were compared to the noncarriers. Hemizygous genotypes for X chromosome genes also qualified for the recessive model.

For the UTR analysis, we used UTRs of all transcript isoforms from ENSEMBL[12] v92 annotation (gtf) and analysed 3 UTR categories – 5'UTR, 3'UTR and UTR combined. We excluded UTR variants that overlapped with any other CDS regions. The CDS regions are a combination of CCDS[20] r22, ENSEMBL[21] v104 and MANE[22] 1.0. We defined UTR QVs according to their MAF and their predicted deleteriousness. Specifically, variants with a CADD score greater than 5 were classified as deleterious. We analysed the UTR QVs using six distinct UTR-only collapsing models, plus two additional models that combine CDS and UTR QVs (protein-truncating variants (ptv) from CDS, alongside UTR variants with varied MAF cutoff) (Table S11). Median lengths of 5'UTRs, 3'UTRs and CDS regions are 288bp, 1064bp, and 1354bp respectively. The number of variants per UTR depends on the models, with a range of 17 to 169 for 5' UTRs and 68 to 533 for the 3' UTRs (Figure S11).

##### Pan-ancestry meta-analysis

We combined the ancestry specific PheWAS results from each of the 5 ancestries with at least 5 cases in a meta-analysis framework. For binary traits we used our previously described approach[10] applying a Cochran-Mantel-Haenzel (CMH) test to generate combined 2 × 2 × N stratified *p* values, with N representing up to all five genetic ancestry groups. For quantitative traits, we implemented an inverse-variance meta-analysis combining the linear regression results across the 5 ancestries.

##### Gene caution lists

We created dummy phenotypes to correspond to each of the six exome sequence delivery batches for the WES data, and each of the three sequence sites for the WGS data to identify and exclude from analyses genes that reflected effects of sequencing batch (WES)/sequencing site (WGS). The combined list of 61 genes associated ($p \leq 1x10-7$) with either sequencing batch or sequence site, within or across ancestries were removed from all analyses (Table S16).

#### Meta-analysis

We meta-analyzed GWAS summary statistics from five ancestries for 68 quantitative traits and 228 ICD-10 disease outcomes with cases ≥200 participants in each ancestry, representing a total of 482,329 UK Biobank participants. We performed the fixed-effects meta-analysis using the Metal software (released on 2011-03-25) and the inverse-variance weighted method. We performed the heterogeneity analysis and used the $I^2$ statistic to identify variants that have different effect sizes across populations. To define genome-wide significant loci for each trait, we first extracted all genome-wide significant variants significant ($P \leq 5$ x 10-8) and the flanking region (±500 Kb) around each variant, we then iteratively merged all regions until no overlapping regions remained. The most significant variant in each merged region was defined as the sentinel variant. The whole MHC region (chr6:25.5–34.0Mb) was treated as a single genomic region.

#### Association testing for structural variants

We tested for association with quantitative traits based on the linear mixed model implemented in BOLT-LMM[23]. We used BOLT-LMM to calculate leave-one-chromosome out (LOCO) residuals which we then tested for association using simple linear regression. We used logistic regression to test for the association between sequence variants and binary traits. We tested variants for association under the additive model using the expected allele counts as a covariate for quantitative traits and integrating over the possible genotypes for binary traits. Sequence center (Vanguard, Sanger, deCODE), other available individual characteristics that correlate with the trait were additionally included in the model; sex, age, and principal components in order to adjust for population stratification. Association analyses in cohorts with sample sizes <10,000 were done with linear regression directly instead of BOLT-LMM. The correction factor employed was the intercept of each regression analysis.

We used LD score regression to account for distribution inflation in the dataset due to cryptic relatedness and population stratification [24]. Using 1.1 million variants, we regressed the $\chi 2$ statistics from our GWASs against LD score and used the intercepts as a correction factor. Effect sizes based on the LOCO residuals are shrunk and we rescaled them based on the shrinkage of the 1.1 million variants used in the LD score regression.

#### Code availability

We used publicly available software (URLs are listed below) in conjunction with the above-described algorithms. BamQC (v 1.0.0), https://github.com/DecodeGenetics/BamQC. GraphTyper (v2.7.5), https://github.com/DecodeGenetics/graphtyper. GATK resource bundle (v4.0.12), gs://genomics-public-data/resources/broad/hg38/v0. Svimmer (v0.1), https://github.com/DecodeGenetics/svimmer.
Dipcall (v0.1), https://github.com/lh3/dipcall. RTG Tools (v3.8.4), https://github.com/RealTimeGenomics/rtg-tools. bcl2fastq (v2.20.0.422), https://support.illumina.com/sequencing/sequencing_software/bcl2fastq-conversion-software.html.
Samtools (v1.9), http://www.htslib.org/. Samblaster (v0.1.24) https://github.com/GregoryFaust/samblaster. biobambam2 (v2.0.79), https://github.com/gt1/biobambam2. bambi (v0.11.1, 0.11.2, 0.12.0, 0.12.1, 0.12.2, 0.13.1, 0.14.0), https://github.com/wtsi-npg/bambi. minimap2 (v2.10), https://github.com/lh3/minimap2. We used R (v3.6.0) https://www.r-project.org/ extensively to analyze data and create plots.
Functionally equivalent implementations of analysis workflows can be accessed on the Velsera Seven Bridges Platform:
https://igor.sbgenomics.com/public/apps/admin/sbg-public-data/functional-equivalence-wgs-cwl1-0, https://igor.sbgenomics.com/public/apps/admin/sbg-public-data/gatk-pre-processing-for-variant-discovery-4-2-0-0, https://igor.sbgenomics.com/public/apps/admin/sbg-public-data/gatk-generic-germline-short-variant-per-sample-calling-4-2-0-0.

### Supplementary material:

#### Supplementary Figures

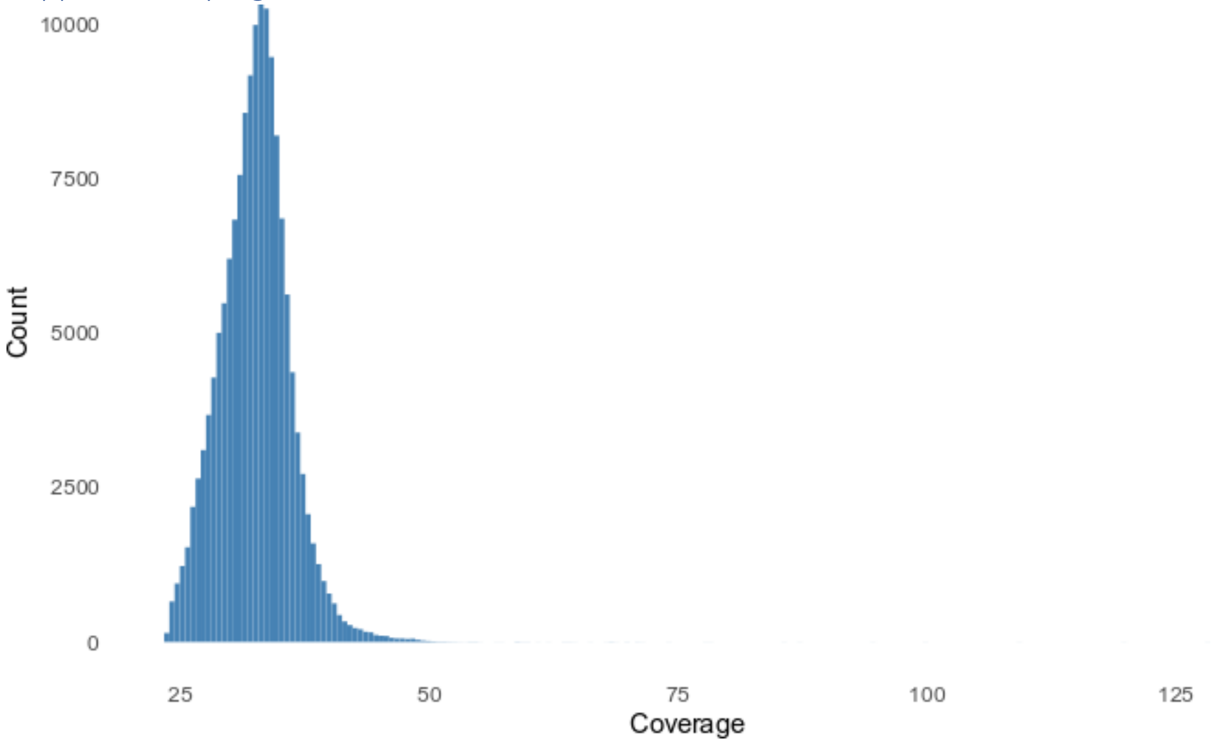


*Fig. S 1 Histogram of average sequence coverage per sample in a subset of 1000 samples.*

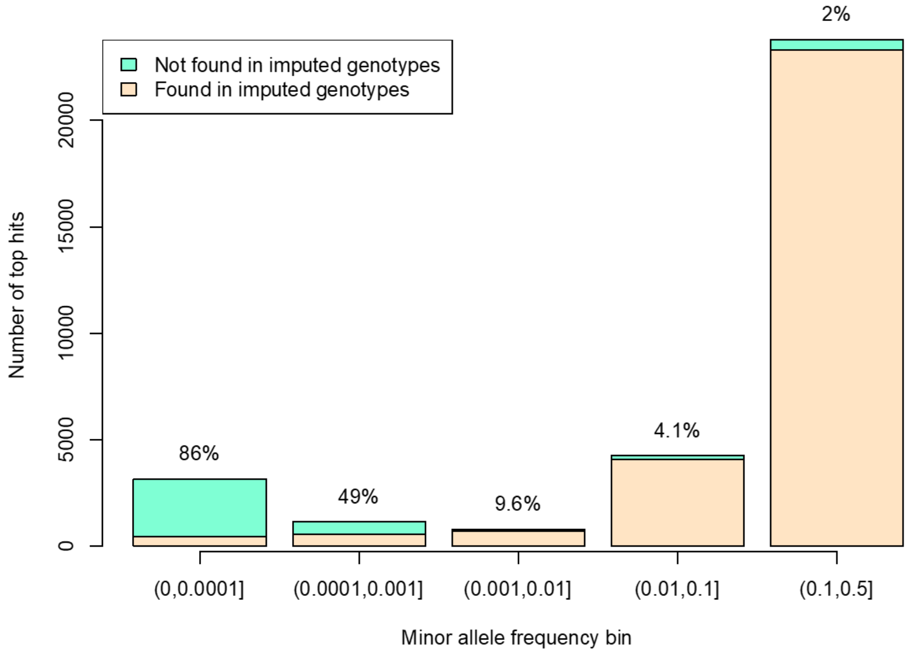


*Fig. S 2 Number of novel associated loci from WGS GWAS compared to those using a set of well-imputed variants from the array data. Each bar represents a different minor allele frequency bin of the lead variant in the locus. The percentages and colors represent the proportion of top hits in each bin that are only seen in WGS.*

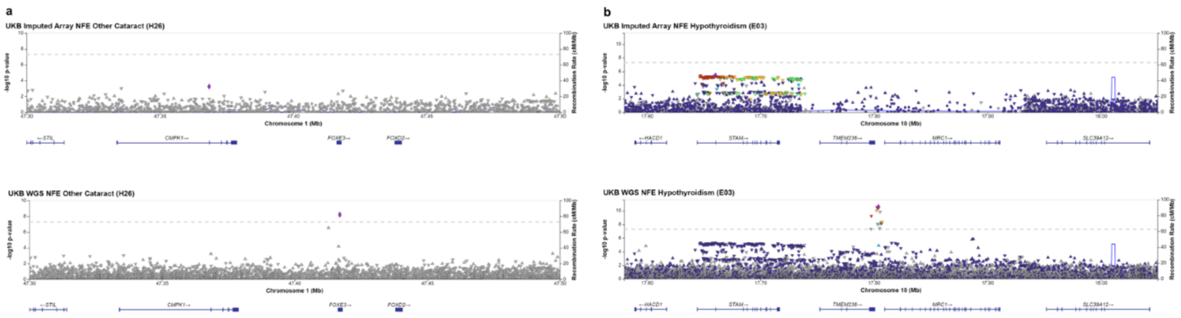


*Fig. S 3 Examples of new WGS top hits: a) legend for example a)? b) A rare frameshift variant (MAF = 5.13-05) in FOXE3 1:47417015:GC:G is found to be significantly associated with the phenotype "other cataract" (H26), p=6.17e-9. The link between FOXE3 and cataract and other ocular diseases was reported in previous familial studies and human and mouse disease models (e.g. Bremond-Gignac et al 2010), but the association was not observed in the UKB imputed array and meta-analysis that included UKB imputed.*

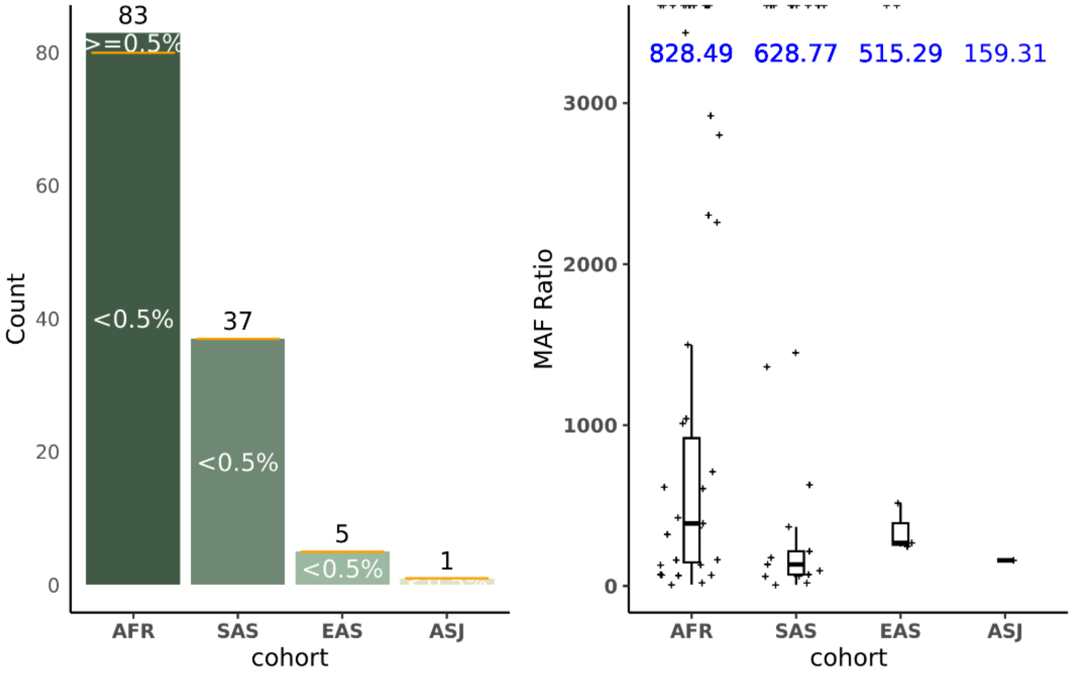


*Fig. S 4 meta-GWS (genome-wide significant) loci driven by non-NFE ancestries. a) among the strongest non-NFE signals, most had NFE MAF (minor allele frequency) <0.5%. b) boxplot of MAFpop/MAFNFE across ancestries. Blue color text shows the median[MAFpop/MAFNFE]. NFE: non-Finnish European; AFR: African; SAS: South Asian; EAS: East Asian; ASJ: Ashkenazi Jewish.*

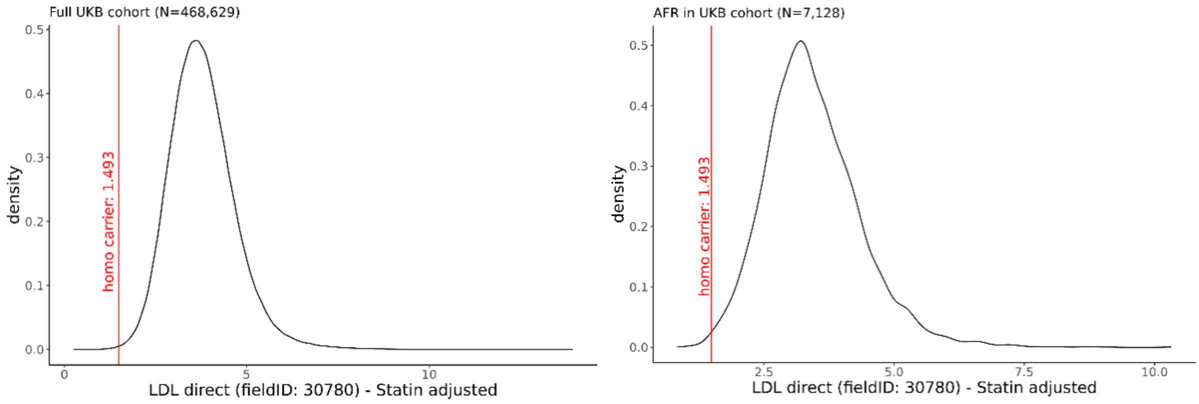


*Fig. S 5 The plasma LDL level of homozygous carrier of PCSK9 Loss-of-function mutation (C679X) in a) UKB full and b) AFR cohort. LDL (mmol/L) is obtained from UKB data field 30780, measured at baseline (initial assessment visit) and adjusted for statin taking at baseline[25].*

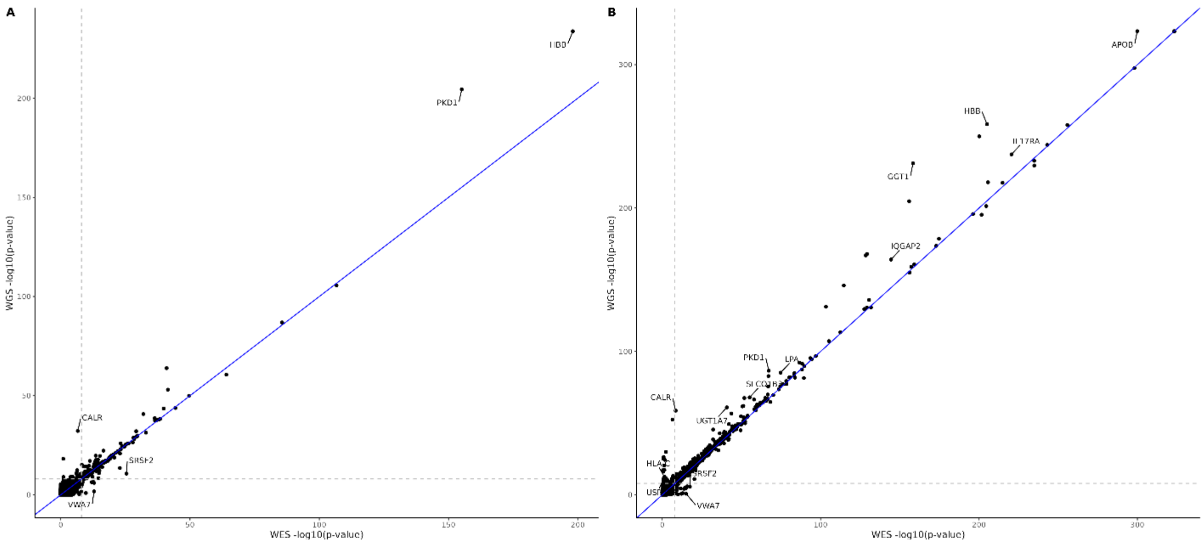


*Fig. S 6 Comparison of rare variant association -log10 meta-analysis p-values between the WES and WGS results for A) binary traits and B) quantitative traits. Associations with absolute difference in Phred scores ≥ 100 are annotated for the most significant phenotype per gene in the WES results.*

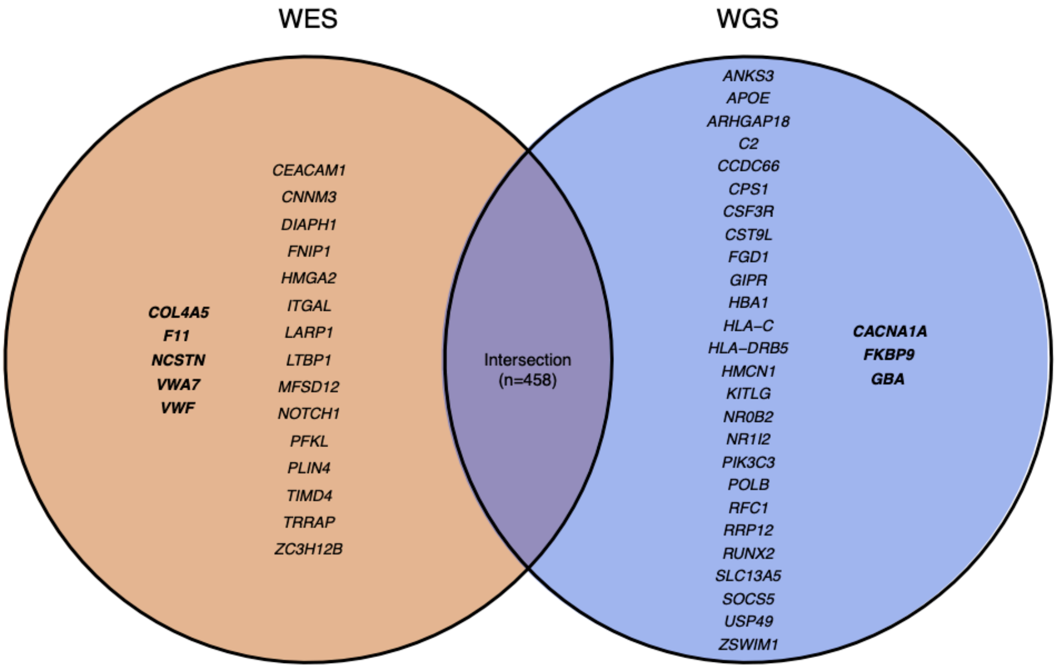


*Fig. S 7 Venn diagram showing genes with significant (p ≤ 1x10-8) phenotype associations across both binary and quantitative traits identified using only one technology (WES or WGS). Genes associated with binary phenotypes are shown in bold.*

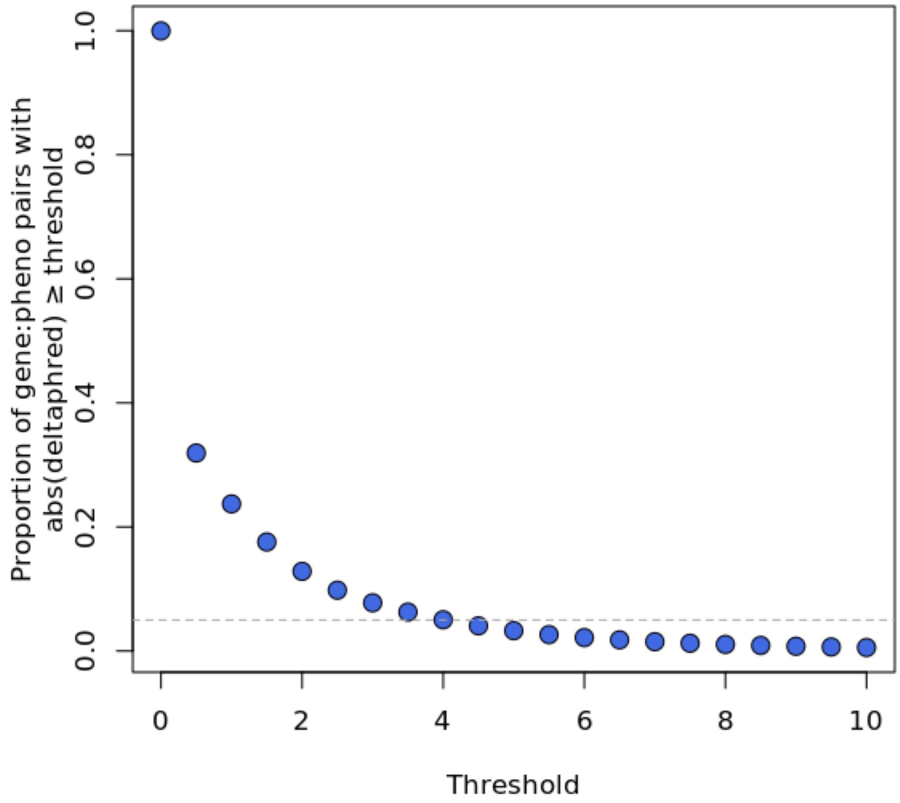


*Fig. S 8 Proportion of gene-phenotype pairs with absolute difference in Phred scores (-10*log10[p-values]) between the WES and WGS results above varying thresholds. Dashed line corresponds to 5%.*

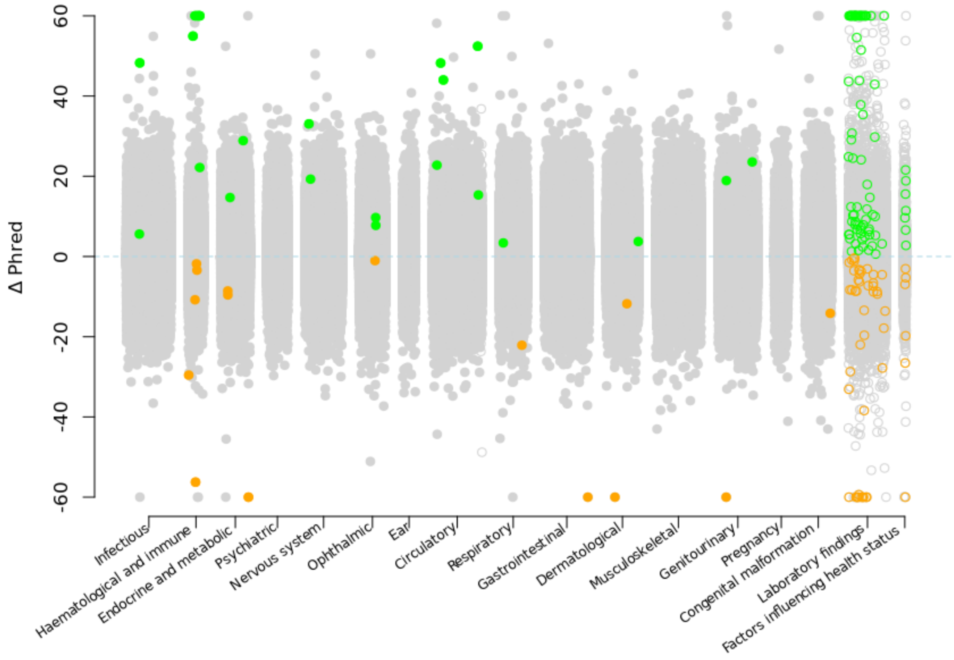


*Fig. S 9 The change in Phred scores between the WGS and WES analyses for 12,963,003 binary genotype-phenotype associations (filled circle) and 1,167,322 quantitative associations (empty circle) stratified by chapter. For gene–phenotype associations that appear in multiple collapsing models, we display only those with the lowest P value within each dataset. The green circles indicate associations that were not significant in the WES analysis but were significant in the WGS analysis. The orange dots represent associations that were originally significant in the WES analysis but became not significant in the WGS analysis. The y axis is capped at ΔPhred = 60 (and -60), equivalent to a P value change of 0.000001.*

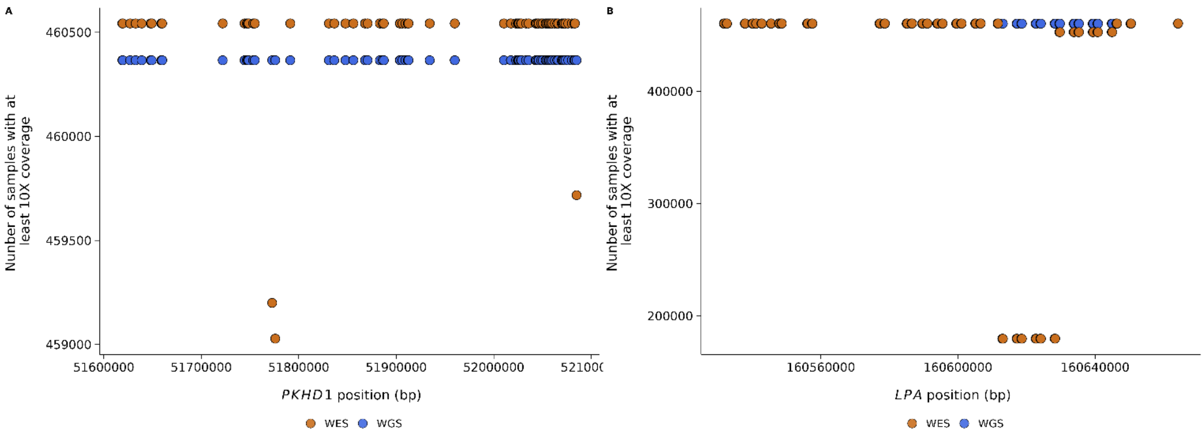


*Fig. S 10 Number of samples with at least 10X coverage across CDS sites in the WES and WGS data for PKHD1 and LPA.*

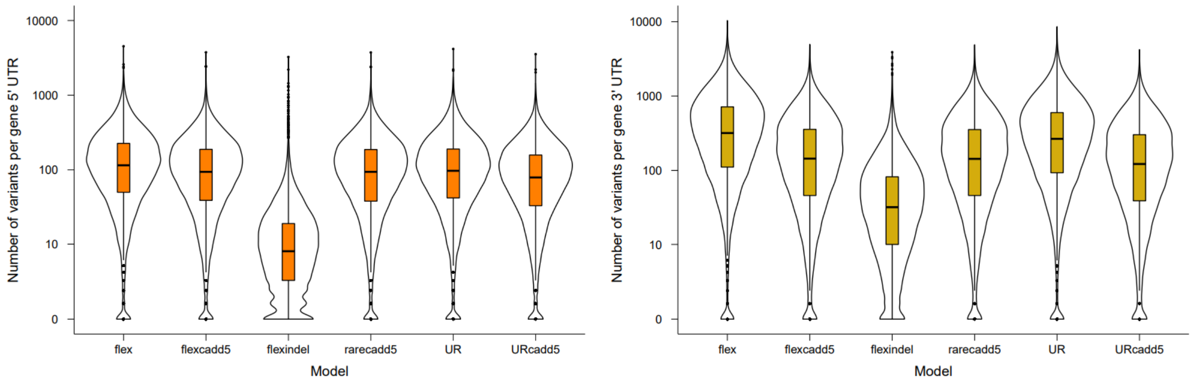


*Fig. S 11 Violin plots showing distribution of number of qualifying variants in 5'UTRs and 3'UTRs according to the six different models.*

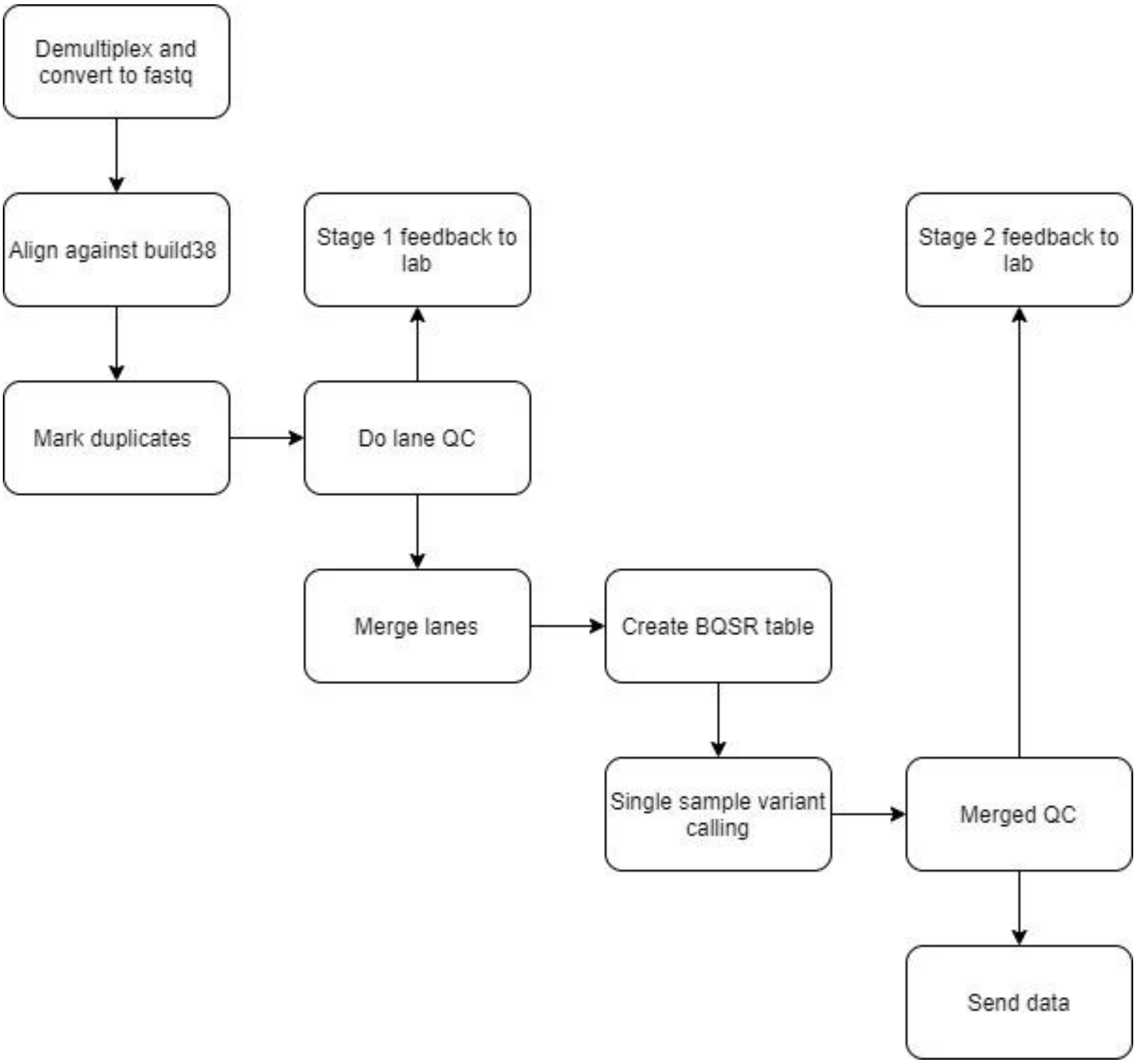


*Fig. S 12 1 Process outline for UKB sequencing pipeline at deCODE genetics.*

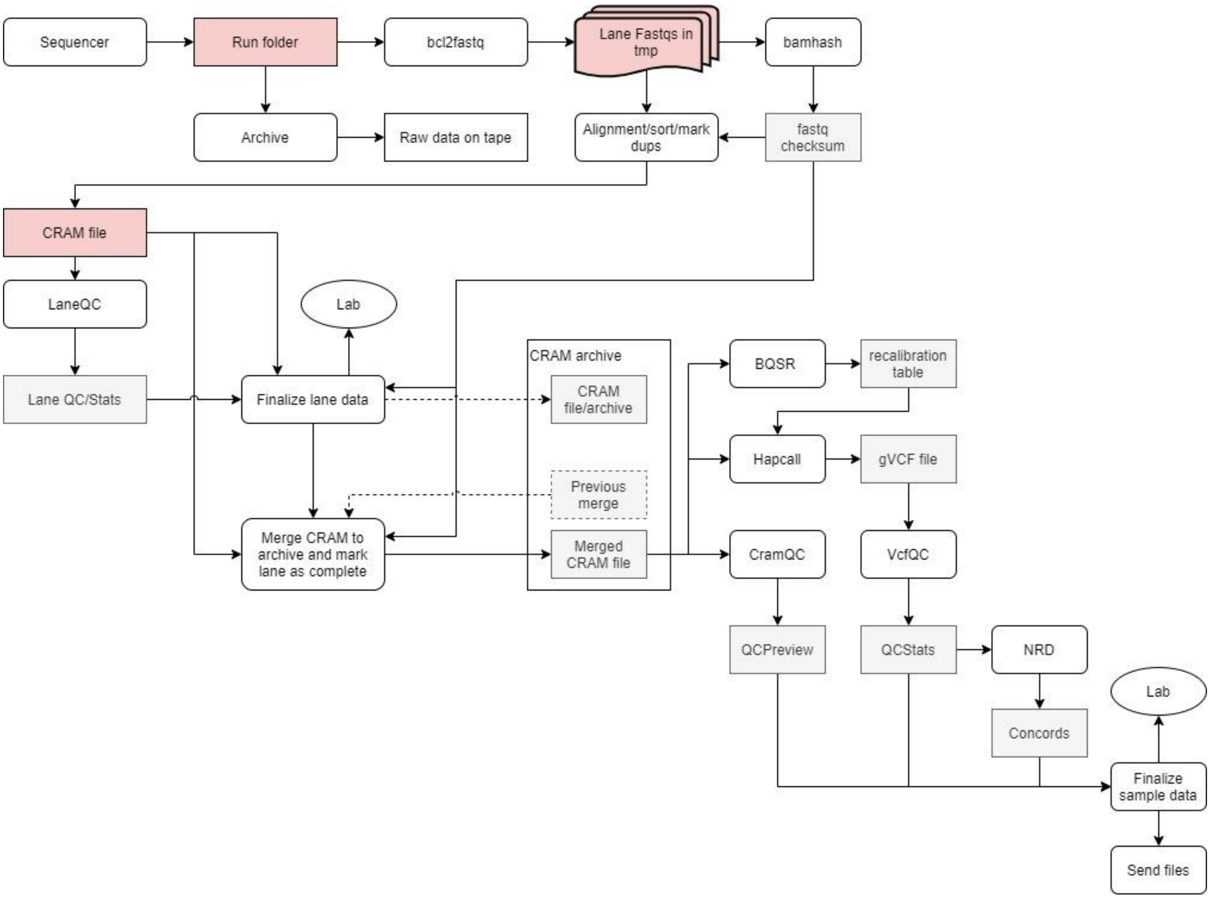


*Fig. S 13 Pipeline for processing of sequence data at deCODE genetics.*

```
QC_VERDICT = 'PASS'

    if freemix_percentage >= 1.0:
        QC_VERDICT = 'REVIEW'

    if coverage < 26:
        QC_VERDICT = 'REVIEW'

    if freemix_percentage >= 5.0:
        QC_VERDICT = 'FAIL'

    if prc_proper_pairs < 95.0:
        QC_VERDICT = 'FAIL'

    if prc_auto_ge_15x < 95.0:
        QC_VERDICT = 'FAIL'

    if discordance_prc is not -1 and discordance_prc >= 2.0:
        QC_VERDICT = 'FAIL'
```

*Fig. S 14 Logic used to compute PASS/FAIL for WGS cram file.*

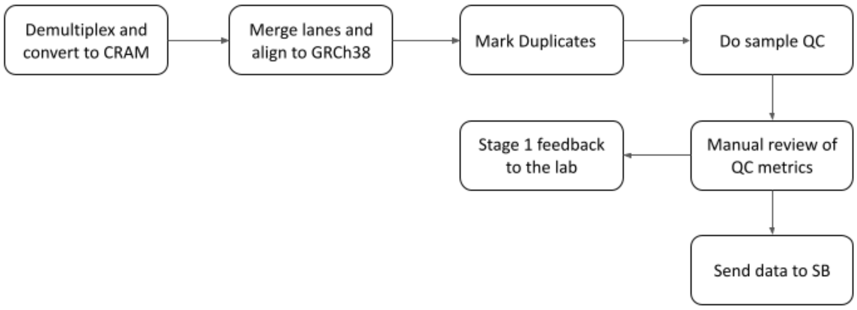


*Fig. S 15 Process outline for Sanger Vanguard sequence data at the Sanger.*

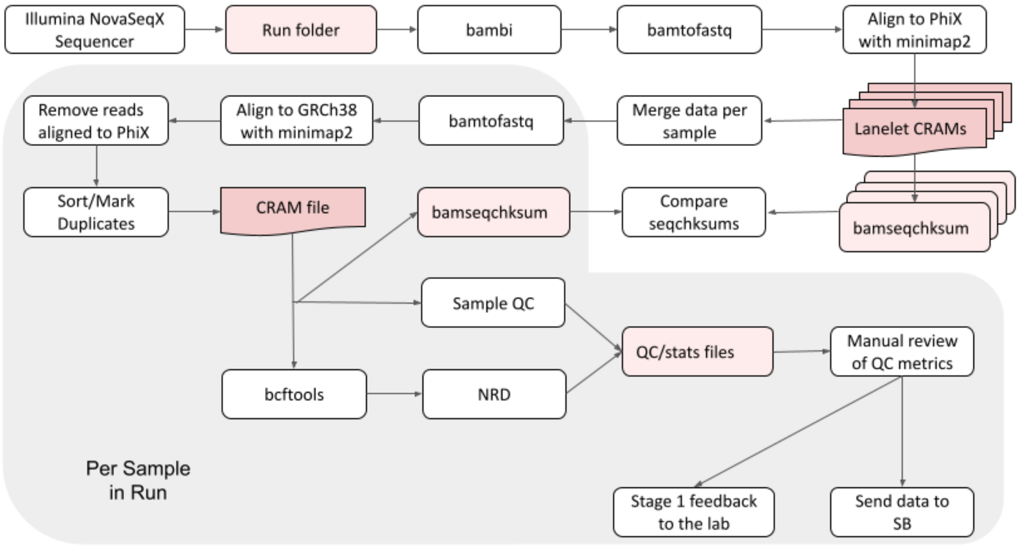


*Fig. S 16 Sanger Vanguard Pipeline for the processing of sequence data at the Sanger.*

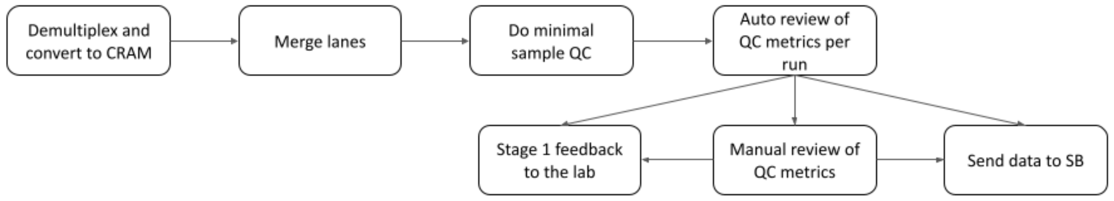


*Fig. S 17 Process outline for Sanger Main Phase sequence data at the Sanger.*

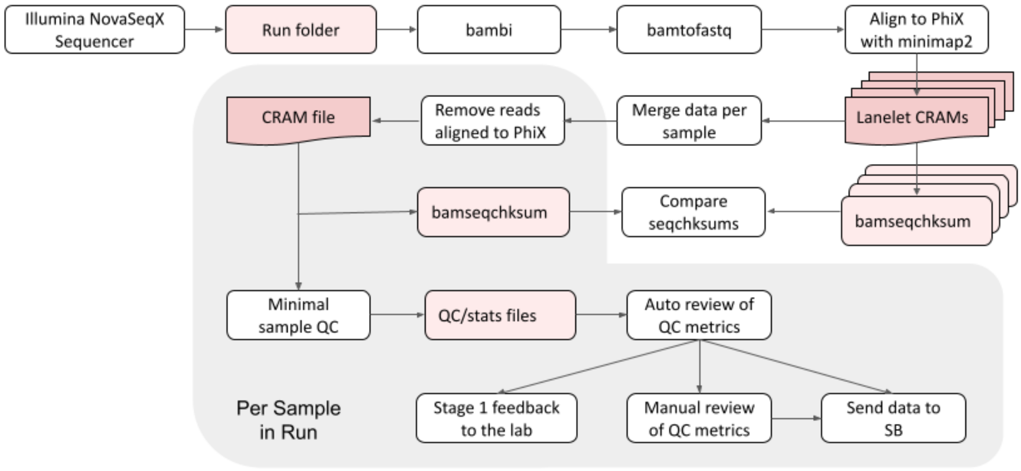


*Fig. S 18 Sanger Main Phase Pipeline for the processing of sequence data at the Sanger.*

### Supplementary Tables

| | Annotation | WGS | WES | Intersection | Unique to WES | Present WES (%) | Missing WES (%) | Present WGS (%) | Missing WGS (%) | Union |
|---|---|---|---|---|---|---|---|---|---|---|
| **SNPs+ Indels** | **Coding** | 12,563,849 | 10,997,033 | 10,813,189 | 183,844 | 86.267 | 13.733 | 98.558 | 1.442 | 12,747,693 |
| | **Splice** | 922,111 | 799,114 | 784,865 | 14,249 | 85.343 | 14.657 | 98.478 | 1.522 | 936,360 |
| | **5' UTR** | 3,127,742 | 973,615 | 944,458 | 29,157 | 30.841 | 69.159 | 99.076 | 0.924 | 3,156,899 |
| | **3' UTR** | 13,941,989 | 1,406,375 | 1,366,180 | 40,195 | 10.058 | 89.942 | 99.713 | 0.287 | 13,982,184 |
| | **Proximal** | 490,613,217 | 12,482,022 | 11,988,515 | 493,507 | 2.542 | 97.458 | 99.900 | 0.100 | 491,106,724 |
| | **Intergenic** | 601,209,600 | 182,763 | 165,217 | 17,546 | 0.030 | 99.970 | 99.997 | 0.003 | 601,227,146 |
| | **Sum** | 1,122,378,508 | 26,840,922 | 26,062,424 | 778,498 | 2.390 | 97.610 | 99.931 | 0.069 | 1,123,157,006 |
| **SNPs** | **Coding** | 11,948,179 | 10,473,822 | 10,325,688 | 148,134 | 86.587 | 13.413 | 98.775 | 1.225 | 12,096,313 |
| | **Splice** | 836,897 | 726,506 | 716,881 | 9,625 | 85.822 | 14.178 | 98.863 | 1.137 | 846,522 |
| | **5' UTR** | 2,864,262 | 897,070 | 874,839 | 22,231 | 31.078 | 68.922 | 99.230 | 0.770 | 2,886,493 |
| | **3' UTR** | 12,388,104 | 1,266,661 | 1,237,013 | 29,648 | 10.200 | 89.800 | 99.761 | 0.239 | 12,417,752 |
| | **Proximal** | 445,380,113 | 11,347,679 | 10,962,189 | 385,490 | 2.546 | 97.454 | 99.914 | 0.086 | 445,765,603 |
| | **Intergenic** | 551,770,600 | 170,187 | 154,649 | 15,538 | 0.031 | 99.969 | 99.997 | 0.003 | 551,786,138 |
| | **Sum** | 1,025,188,155 | 24,881,925 | 24,271,259 | 610,666 | 2.426 | 97.574 | 99.940 | 0.060 | 1,025,798,821 |
| **Indels** | **Coding** | 615,670 | 523,211 | 487,501 | 35,710 | 80.323 | 19.677 | 94.518 | 5.482 | 651,380 |
| | **Splice** | 85,214 | 72,608 | 67,984 | 4,624 | 80.821 | 19.179 | 94.853 | 5.147 | 89,838 |
| | **5' UTR** | 263,480 | 76,545 | 69,619 | 6,926 | 28.307 | 71.693 | 97.439 | 2.561 | 270,406 |
| | **3' UTR** | 1,553,885 | 139,714 | 129,167 | 10,547 | 8.931 | 91.069 | 99.326 | 0.674 | 1,564,432 |
| | **Proximal** | 45,233,104 | 1,134,343 | 1,026,326 | 108,017 | 2.502 | 97.498 | 99.762 | 0.238 | 45,341,121 |
| | **Intergenic** | 49,439,000 | 12,576 | 10,568 | 2,008 | 0.025 | 99.975 | 99.996 | 0.004 | 49,441,008 |
| | **Sum** | 97,190,353 | 1,958,997 | 1,791,165 | 167,832 | 2.012 | 97.988 | 99.828 | 0.172 | 97,358,185 |

*Table S 1 Comparison of the number of SNP and Indel variants discovered in this study using the GraphTyper dataset, and the number of variants discovered through WES of the UKB, split by functional annotation. Data is shown for the number of SNPs and indels combined and separately.*

a)

| | Annotation | WGS | WES | Intersection | Unique to WES | Present WES (%) | Missing WES (%) | Present WGS (%) | Missing WGS (%) | Union |
|---|---|---|---|---|---|---|---|---|---|---|
| **SNPs+ Indels** | **Coding** | 12,563,849 | 10,997,033 | 10,813,189 | 183,844 | 86.267 | 13.733 | 98.558 | 1.442 | 12,747,693 |
| | **Splice** | 922,111 | 799,114 | 784,865 | 14,249 | 85.343 | 14.657 | 98.478 | 1.522 | 936,360 |
| | **5' UTR** | 3,127,742 | 973,615 | 944,458 | 29,157 | 30.841 | 69.159 | 99.076 | 0.924 | 3,156,899 |
| | **3' UTR** | 13,941,989 | 1,406,375 | 1,366,180 | 40,195 | 10.058 | 89.942 | 99.713 | 0.287 | 13,982,184 |
| | **Proximal** | 490,613,217 | 12,482,022 | 11,988,515 | 493,507 | 2.542 | 97.458 | 99.900 | 0.100 | 491,106,724 |
| | **Intergenic** | 601,209,600 | 182,763 | 165,217 | 17,546 | 0.030 | 99.970 | 99.997 | 0.003 | 601,227,146 |
| | **Sum** | 1,122,378,508 | 26,840,922 | 26,062,424 | 778,498 | 2.390 | 97.610 | 99.931 | 0.069 | 1,123,157,006 |
| **SNPs** | **Coding** | 11,948,179 | 10,473,822 | 10,325,688 | 148,134 | 86.587 | 13.413 | 98.775 | 1.225 | 12,096,313 |
| | **Splice** | 836,897 | 726,506 | 716,881 | 9,625 | 85.822 | 14.178 | 98.863 | 1.137 | 846,522 |
| | **5' UTR** | 2,864,262 | 897,070 | 874,839 | 22,231 | 31.078 | 68.922 | 99.230 | 0.770 | 2,886,493 |
| | **3' UTR** | 12,388,104 | 1,266,661 | 1,237,013 | 29,648 | 10.200 | 89.800 | 99.761 | 0.239 | 12,417,752 |
| | **Proximal** | 445,380,113 | 11,347,679 | 10,962,189 | 385,490 | 2.546 | 97.454 | 99.914 | 0.086 | 445,765,603 |
| | **Intergenic** | 551,770,600 | 170,187 | 154,649 | 15,538 | 0.031 | 99.969 | 99.997 | 0.003 | 551,786,138 |
| | **Sum** | 1,025,188,155 | 24,881,925 | 24,271,259 | 610,666 | 2.426 | 97.574 | 99.940 | 0.060 | 1,025,798,821 |
| **Indels** | **Coding** | 615,670 | 523,211 | 487,501 | 35,710 | 80.323 | 19.677 | 94.518 | 5.482 | 651,380 |
| | **Splice** | 85,214 | 72,608 | 67,984 | 4,624 | 80.821 | 19.179 | 94.853 | 5.147 | 89,838 |
| | **5' UTR** | 263,480 | 76,545 | 69,619 | 6,926 | 28.307 | 71.693 | 97.439 | 2.561 | 270,406 |
| | **3' UTR** | 1,553,885 | 139,714 | 129,167 | 10,547 | 8.931 | 91.069 | 99.326 | 0.674 | 1,564,432 |
| | **Proximal** | 45,233,104 | 1,134,343 | 1,026,326 | 108,017 | 2.502 | 97.498 | 99.762 | 0.238 | 45,341,121 |
| | **Intergenic** | 49,439,000 | 12,576 | 10,568 | 2,008 | 0.025 | 99.975 | 99.996 | 0.004 | 49,441,008 |
| | **Sum** | 97,190,353 | 1,958,997 | 1,791,165 | 167,832 | 2.012 | 97.988 | 99.828 | 0.172 | 97,358,185 |

b)

| Annotation | WGS | TOPMED | Intersection | Unique to TOPMED | Present TOPMED(%) | Missing TOPMED(%) | Present WGS(%) | Missing WGS(%) | Union |
|---|---|---|---|---|---|---|---|---|---|
| **Coding** | 12,563,849 | 8,455,257 | 4,722,443 | 3,732,814 | 51.883 | 48.117 | 77.095 | 22.905 | 16,296,663 |
| **Splice** | 922,111 | 600,367 | 308,565 | 291,802 | 49.457 | 50.543 | 75.962 | 24.038 | 1,213,913 |
| **5' UTR** | 3,127,742 | 2,163,045 | 1,218,870 | 944,175 | 53.121 | 46.879 | 76.813 | 23.187 | 4,071,917 |
| **3' UTR** | 13,941,989 | 9,722,052 | 5,306,755 | 4,415,297 | 52.960 | 47.040 | 75.948 | 24.052 | 18,357,286 |
| **Proximal** | 490,613,217 | 345,799,288 | 187,192,338 | 158,606,950 | 53.264 | 46.736 | 75.570 | 24.430 | 649,220,167 |
| **Intergenic** | 601,209,600 | 442,657,286 | 228,690,756 | 213,966,530 | 54.302 | 45.698 | 73.752 | 26.248 | 815,176,130 |
| **Sum** | 1,122,378,508 | 809,397,295 | 427,439,727 | 381,957,568 | 53.804 | 46.196 | 74.610 | 25.390 | 1,504,336,076 |

c)

| Annotation | TOPMED | GNOMAD | Intersection | Unique to GNOMAD | Present GNOMAD(%) | Missing GNOMAD(%) | Present TOPMED(%) | Missing TOPMED(%) | Union |
|---|---|---|---|---|---|---|---|---|---|
| **Coding** | 8,455,257 | 7,295,558 | 5,347,135 | 1,948,423 | 70.125 | 29.875 | 81.272 | 18.728 | 10,403,680 |
| **Splice** | 600,367 | 547,462 | 366,375 | 181,087 | 70.057 | 29.943 | 76.827 | 23.173 | 781,454 |
| **5' UTR** | 2,163,045 | 1,907,955 | 1,366,899 | 541,056 | 70.558 | 29.442 | 79.991 | 20.009 | 2,704,101 |
| **3' UTR** | 9,722,052 | 8,889,457 | 6,190,106 | 2,699,351 | 71.566 | 28.434 | 78.269 | 21.731 | 12,421,403 |
| **Proximal** | 345,799,288 | 331,085,507 | 221,412,772 | 109,672,735 | 72.675 | 27.325 | 75.904 | 24.096 | 455,572,023 |
| **Intergenic** | 442,657,286 | 411,385,147 | 273,909,902 | 137,475,245 | 70.912 | 29.088 | 76.303 | 23.697 | 580,132,531 |
| **Sum** | 809,397,295 | 761,111,086 | 508,593,189 | 252,517,897 | 71.667 | 28.333 | 76.213 | 23.787 | 1,062,015,192 |

*Table S 2 Comparison of the number of variants discovered in this study using the GraphTyper dataset, TopMed and Gnomad, split by functional annotation. A) This study compared to TOPMED. B) This study compared to Gnomad. C) Gnomad compared to TOPMED.*

| variant metrics | whole genome | autosome | chrX | chrY | chrM | alt contigs |
|---|---|---|---|---|---|---|
| **variant sites** | 1,109,854,569 | 1,013,260,852 | 45,587,922 | 6,215,922 | 16,529 | 44,773,344 |
| **variant alleles** | 1,494,611,198 | 1,340,689,096 | 59,458,821 | 9,466,466 | 67,312 | 84,929,503 |
| **SNV** | 1,289,650,789 | 1,157,537,506 | 51,840,675 | 8,185,380 | 41,230 | 72,045,998 |
| **INDEL** | 204,960,409 | 648,648,544 | 28,402,615 | 3,279,332 | 16,210 | 27,044,612 |
| **transition** | 707,391,313 | 508,888,962 | 23,438,060 | 4,906,048 | 25,020 | 45,001,386 |
| **transversion** | 582,259,476 | 183,151,590 | 7,618,146 | 1,281,086 | 26,082 | 12,883,505 |
| **insertion** | 108,305,453 | 96,600,630 | 4,060,050 | 591,019 | 22,307 | 7,031,447 |
| **deletion** | 96,654,956 | 86,550,960 | 3,558,096 | 690,067 | 3,775 | 5,852,058 |

*Table S 3 Summary statistics of variants called in 490,541 individuals in the DRAGEN aggregate dataset. Numbers are after applying DRAGEN Machine Learning recalibration cutoff QUAL>=3*

A) SNP+Indel

| GIAB sample | #Variants | Sensitivity | Precision | F1-score |
|---|---|---|---|---|
| HG001 | 3,376,982 | 98.05% | 98.21% | 98.13% |
| HG002 | 3,434,822 | 98.00% | 98.31% | 98.16% |
| HG003 | 3,283,999 | 97.91% | 98.12% | 98.02% |
| HG004 | 3,322,079 | 98.01% | 98.30% | 98.15% |
| HG005 | 3,234,416 | 97.78% | 98.66% | 98.22% |
| HG006 | 3,325,801 | 98.17% | 98.27% | 98.22% |
| HG007 | 3,346,308 | 98.20% | 98.32% | 98.26% |
| Average | 3,332,058 | 98.02% | 98.31% | 98.17% |

B) SNP

| GIAB sample | #Variants | Sensitivity | Precision | F1-score |
|---|---|---|---|---|
| HG001 | 2,909,899 | 98.29% | 98.34% | 98.32% |
| HG002 | 2,973,969 | 98.17% | 98.39% | 98.28% |
| HG003 | 2,829,271 | 98.15% | 98.24% | 98.19% |
| HG004 | 2,856,376 | 98.22% | 98.38% | 98.30% |
| HG005 | 2,841,834 | 97.85% | 98.74% | 98.29% |
| HG006 | 2,929,927 | 98.24% | 98.28% | 98.26% |
| HG007 | 2,948,112 | 98.27% | 98.32% | 98.29% |
| Average | 2,898,484 | 98.17% | 98.38% | 98.28% |

C) Indel

| GIAB sample | #Variants | Sensitivity | Precision | F1-score |
|---|---|---|---|---|
| HG001 | 467,083 | 96.58% | 97.41% | 96.99% |
| HG002 | 460,853 | 96.94% | 97.85% | 97.39% |
| HG003 | 454,728 | 96.49% | 97.35% | 96.92% |
| HG004 | 465,703 | 96.78% | 97.79% | 97.28% |
| HG005 | 392,582 | 97.27% | 98.09% | 97.68% |
| HG006 | 395,874 | 97.62% | 98.26% | 97.94% |
| HG007 | 398,196 | 97.70% | 98.32% | 98.01% |
| Average | 433,574 | 97.05% | 97.87% | 97.46% |

*Table S 4 Genome in a bottle (GIAB) v3.3.2 truth set comparison of GraphTyper variant calls. Calls of each sample were extracted from the full set of variant calls. F1-score is the harmonic mean of Sensitivity and Precision. A) all variant types, B) SNPs only C) indels only.*

| GT rate | # sites | % sites |
|---|---|---|
| **0%-10%** | 49,251,611 | 4.44% |
| **10%-20%** | 5,361,149 | 0.48% |
| **20%-30%** | 4,739,631 | 0.43% |
| **30%-40%** | 4,688,034 | 0.42% |
| **40%-50%** | 9,538,084 | 0.86% |
| **50%-60%** | 4,881,545 | 0.44% |
| **60%-70%** | 5,365,681 | 0.48% |
| **70%-80%** | 6,801,069 | 0.61% |
| **80%-90%** | 9,120,448 | 0.82% |
| **90%-100%** | 1,010,107,317 | 91.01% |

*Table S 5 Site level genotyping rate in the DRAGEN aggregate dataset. Genotyping (GT) rate is a metric to assess the quality of genotyping. For common variants, typical cutoff is 90% whereas for rare variants the cutoff can be between 10%-90%, depending on target sensitivity.*

Table S6. GWAS_phenotypes_metadata – See separate Excel file.

| Phenotypes | WGS GWAS variants | | Array imputed variants | |
|---|---|---|---|---|
| | Total top hits | Novel top hits in WGS | Total top hits | New WGS lead variant |
| 763 binary disease traits | 8,132 | 2,872 | 5,283 | 492 |
| 71 quantitative biomarker traits | 24,991 | 1,119 | 24,074 | 2,492 |

*Table S7. Number of total and novel top hits identified from the WGS GWAS and those with only well-imputed array variants.*

Table S8. Trans-ancestry meta-GWAS results for a) 68 quantitative traits and b) 228 ICD-10 disease outcomes. – See separate Excel file.

Table S9. Associations with sentinel variants found significant only in non-NFE ancestries. – See separate Excel file.

Table S10. UKB WGS revealed heterozygous and homozygous carriers of pLoF/P/LP variants in the 81 ACMG genes. – See separate Excel file.

Table S11. Phenotypes included in region-based collapsing analysis PheWASs. – See separate Excel file.

| COLLAPSING MODEL | GNOMAD MAF* | UKB MAF | UKB COHORT NO CALL OR QC FAIL^ | VARIANT TYPE | REVEL, MTR AND CADD CUT-OFFS |
|---|---|---|---|---|---|
| CDS PheWAS | | | | | |
| syn (synonymous negative control) | **≤ 0.005%** | **≤ 0.05%** | **≤ 0.005%** | **Synonymous** | **-** |
| ptv (Protein Truncating) | **≤ 0.1% (popmax)$^{+}$** | **≤ 0.1%** | **≤ 0.01%** | **PTV** | **-** |
| ptv5pct (Protein Truncating; ≤ 5% MAF) | **≤ 5% (popmax)** | **≤ 5%** | **≤ 0.5%** | **PTV** | **-** |
| UR (Ultra-rare damaging) | **0%** | **≤ 0.005%** | **≤ 0.001%** | **Non-synonymous** | **REVEL ≥ 0.25** |
| URmtr (Ultra-rare damaging, MTR informed) | **0%** | **≤ 0.005%** | **≤ 0.001%** | **Non-synonymous** | **REVEL ≥ 0.25 MTR ≤ 25$^{th}$ %ile or intragenic MTR ≤ 50$^{th}$ %ile** |
| raredmg (Rare damaging) | **≤ 0.005%** | **≤ 0.025%** | **≤ 0.005%** | **Missense** | **REVEL ≥ 0.25** |
| raredmgmtr (Rare damaging, MTR informed) | **≤ 0.005%** | **≤ 0.025%** | **≤ 0.005%** | **Missense** | **REVEL ≥ 0.25 MTR ≤ 25$^{th}$ %ile or intragenic MTR ≤ 50$^{th}$ %ile** |
| flexdmg (Flexible MAF, damaging non-synonymous) | **≤ 0.1% (popmax)** | **≤ 0.1%** | **≤ 0.01%** | **Non-synonymous** | **REVEL ≥ 0.25** |
| flexnonsynmtr (Flexible MAF, non-synonymous, MTR informed) | **≤ 0.1% (popmax)** | **≤ 0.1%** | **≤ 0.01%** | **Non-synonymous** | **MTR ≤ 25$^{th}$ %ile or intragenic MTR ≤ 50$^{th}$ %ile** |
| ptvraredmg (PTV or rare damaging models combined) | **PTV ≤ 0.1% (popmax) missense ≤ 0.005% and ≤ 0.05% (popmax)** | **PTV ≤ 0.1% missense ≤ 0.025%** | **≤ 0.01%** | **Non-synonymous** | **REVEL ≥ 0.25** |
| rec (Non-synonymous recessive) | **≤ 1% (popmax) ≤ 10 homozygous calls** | **≤ 1%** | **≤ 0.1%** | **Non-synonymous** | **-** |
| UTR PheWAS | | | | | |
| UR (Ultra-rare) | **0%** | **≤ 0.002%** | **≤ 0.001%** | **UTR (Non-coding)** | **-** |
| URcadd5 (Ultra-rare with CADD) | **0%** | **≤ 0.002%** | **≤ 0.001%** | **UTR (Non-coding)** | **CADD > 5** |
| Flex (Flexible) | **≤0.1% and ≤ 0.1% (popmax)$^{+}$** | **≤ 0.1%** | **≤ 0.01%** | **UTR (Non-coding)** | **-** |
| flexcadd5 (Flexible with CADD) | **≤0.1% and ≤ 0.1% (popmax)$^{+}$** | **≤ 0.1%** | **≤ 0.01%** | **UTR (Non-coding)** | **CADD >** |
| Flexindel (Flexible INDELs) | **≤0.1% and ≤ 0.1% (popmax)$^{+}$** | **≤ 0.1%** | **≤ 0.01%** | **UTR (Non-coding)** | **-** |
| rarecadd5 (rare with CADD) | **≤ 0.01%** | **≤ 0.025%** | **≤ 0.005%** | **UTR (Non-coding)** | **CADD > 5** |
| CDS + UTR PheWAS | | | | | |
| $PTV_{CDS}$ + $UR_{UTR}$ (PTV or UTR ultra-rare models combined) | **PTV ≤ 0.1% (popmax)$^{+}$ and UR UTR = 0%** | **≤ 0.1% (PTV); ≤ 0.002% (UTR)** | **PTV ≤ 0.01% and UR UTR ≤ 0.001%** | **PTV + UTR** | **-** |
| $PTV_{CDS}$ + $Flex_{UTR}$ (PTV or UTR flexible models combined) | **PTV ≤ 0.1% (popmax)$^{+}$ and Flex UTR ≤0.1% and ≤ 0.1% (popmax)$^{+}$** | **≤ 0.1% (PTV); ≤ 0.1% (UTR)** | **PTV ≤ 0.01% and Flex UTR ≤ 0.01%** | **PTV + UTR** | **-** |

*Table S 12 Genetic models for region-based collapsing analysis PheWASs.*

* reflects the gnomAD global_raw MAF unless otherwise specified.
^ reflects the maximum proportion of UKB exome sequences permitted to either have ≤ 10-fold coverage at variant site or carry a low-confidence variant that did not meet one of the quality-control thresholds applied to collapsing analyses (see methods).
+ The term 'popmax' refers to the gnomAD non-bottlenecked population with the maximum allele frequency
(CDS = coding Sequence; UTR = untranslated region; MAF = minor allele frequency; QC = quality control; MTR = Missense Tolerance Ratio; CADD = Combined Annotation Dependent Depletion score)
Synonymous: synonymous_variant

**PTV**: exon_loss_variant, frameshift_variant, start_lost, stop_gained, stop_lost, splice_acceptor_variant, splice_donor_variant, gene_fusion, bidirectional_gene_fusion, rare_amino_acid_variant, transcript_ablation
**Missense**: missense_variant_splice_region_variant, missense_variant
**Nonsynonymous**: exon_loss_variant, frameshift_variant, start_lost, stop_gained, stop_lost, splice_acceptor_variant, splice_donor_variant, gene_fusion, bidirectional_gene_fusion, rare_amino_acid_variant, transcript_ablation, conservative_inframe_deletion, conservative_inframe_insertion, disruptive_inframe_insertion, disruptive_inframe_deletion, missense_variant_splice_region_variant, missense_variant, protein_altering_variant
**UTR**: 5_prime_UTR_variant, 5_prime_UTR_premature_start_codon_gain_variant, 3_prime_UTR_variant

Table S13. Significant ($p \leq 1x10-8$) gene-phenotype associations identified in the coding PheWAS collapsing analysis across both WES and WGS datasets. – See separate Excel file.

Table S14. Significant ($p \leq 1x10-8$) gene-phenotype associations identified in the UTR PheWAS collapsing analysis across 5′ UTR, 3′ UTR, 5′ + 3′ UTR and CDS + 5′ + 3′ UTR. – See separate Excel file.

| | **$ASJ_p$** | **$AFR_p$** | **$EAS_P$** | **$SAS_p$** | **$NFE_p$** | **$NFE_{OR}$** | **Meta-analysis$_p$** | **Meta-analysis$_{OR}$** |
|---|---|---|---|---|---|---|---|---|
| $CDS_{PTV}$ | 0.003 | 1 | 1 | 0.202 | 0.038 | 1.48 | 0.016 | 1.54 |
| $CDS_{UR}$ | 1 | 1 | 0.195 | 1 | 1 | 0.97 | 1 | 0.97 |
| 5′ $UTR_{UR}$ | 1 | 0.350 | 1 | 0.368 | 0.111 | 1.40 | 0.096 | 1.42 |
| 3′ $UTR_{UR}$ | 1 | 0.001 | 0.039 | 0.044 | $3.02 \times 10^{-5}$ | 1.70 | $2.11 \times 10^{-7}$ | 1.85 |
| 5′ + 3′ $UTR_{UR}$ | 1 | 0.002 | 0.089 | 0.030 | $2.01 \times 10^{-5}$ | 1.61 | $1.65 \times 10^{-7}$ | 1.73 |
| $CDS_{PTV}$ + 5′ + 3′ $UTR_{UR}$ | 0.024 | 0.009 | 0.134 | 0.014 | $2.25 \times 10^{-6}$ | 1.58 | **$9.24 \times 10^{-9}$** | 1.68 |

Table S15: NWD1-Kidney calculus association P-values and OR (NFE and meta-analysis) calculated in different strategies.

| **Allele Frequency** | **0.0001%-0.001%** | **0.001%-0.01%** | **0.01%-0.1%** | **0.1%-1%** | **1%-10%** | **10%-100%** |
|---|---|---|---|---|---|---|
| **Benign/Likely Benign** | 7 | 15 | 22 | 15 | 12 | 30 |
| **Uncertain Signficance** | 64 | 53 | 28 | 8 | 5 | 12 |
| **Pathogenic/Likely Pathogenic** | 132 | 61 | 18 | 2 | 1 | 0 |

Table S16: Number of structural variants that are annotated in ClinVar and found in the current dataset stratified by pathogenicity and number of carriers.

Table S17: Gene level cautions for region-based collapsing analysis PheWASs. Genes identified as being associated ($p \leq 1x10-7$) with WES sequencing batch or WGS sequencing site. – See separate Excel file.

A) Trio

| Method | FDR | TP | #Variants |
|---|---|---|---|
| GraphTyper | 11.90% | 58,065,835 | 65,919,123 |
| GraphTyperHQ | 5.20% | 56,556,228 | 59,650,581 |

B) Twin consistency table

| Method | ICPM | Non-ref consistency | Number of non-ref calls |
|---|---|---|---|
| GraphTyper | 89.6 | 94.50% | 865,149,777 |
| GraphTyperHQ | 22.2 | 98.48% | 771,842,751 |

Table S18 A) Estimate of false discovery rate (FDR) and number of true positive (TP) variants among the 1,045 parent-offspring trios. The estimates are determined from the allele transmission ratios from parent to offspring. B) Genotype consistency across among the 177 monozygotic twin pairs. ICPM = number of inconsistent genotypes per 1Mb.

| Parameter | Information Requested | Definition |
|---|---|---|
| prc_auto_ge_15x | Coverage | PCT_15X from .wgsmetrics_autosome in QCPreview |
| Coverage | autosomal mean coverage | MEAN_COVERAGE * (1.0 - PCT_EXC_DUPE - PCT_EXC_OVERLAP - PCT_EXC_ADAPTER) / (1.0 - PCT_EXC_TOTAL) from .wgsmetrics_autosome in QCPreview |
| genetic_sex | Sex | if NX<=0.3 then "Female" else if NX>=0.7 then "Male" else "Undetermined" from .sexcheck output file in QCStats |
| Yield | Yield | GENOME_TERRITORY * MEAN_COVERAGE * (1.0 - PCT_EXC_DUPE - PCT_EXC_OVERLAP - PCT_EXC_ADAPTER) / (1.0 - PCT_EXC_TOTAL) from .wgsmetrics output file in QCPreview |
| read_haps_error_percentage | Read_haps | 100*DOUBLE_ERROR_FRACTION from .contamination output file in QCStats |
| freemix_percentage | Freemix/Verify Bam ID | 100 * FREEMIX from .verifyBamId.selfSM output file in QCStats |
| prc_proper_pairs | Proportion of mapped read pairs | 100 * (reads_properly_paired/reads_mapped) from .stats output file in QCPreview |
| discordance_prc | NRD Genotyping | 100 * (1.0 - NON_REF_GENOTYPE_CONCORDANCE) from .genotype_concordance_summary_metrics in Concords or -1 if chip genotypes are not available |

Table S19. QA/QC metrics derived from the files delivered to the UKB.
The result is written to a file, qaqc_metric.

| Column | Min | Max | Flag | Explanation |
|---|---|---|---|---|
| SAMPLE_ID | | | | Read group ID |
| LANE | | | | Lane ID (=Read group ID) |
| FAILURE_FLAGS | | | | Failure flag |
| JOINT_CALLING_FLAGS | | | | Joint calling failure flag |
| STRICT_FLAGS | | | | Strict failure flag |
| TOTAL_BPS | 3e8 | 1e14 | C | Total basepairs |
| TOTAL_READ_PAIRS | | | | Total read pairs |
| READ_LENGTH | | | | Read length |
| MEAN_BASE_QUAL_PER_READ | 30 | 100 | Q | Mean of base calling quality |
| STD_BASE_QUAL_PER_READ | -1 | 10 | Q | Std dev of mean base calling quality |
| MEAN_N_COUNT_PER_READ | -1 | 10 | N | Mean Percentage N |
| STD_N_COUNT_PER_READ | -1 | 30 | N | Std dev of Percentage N |
| MEAN_GC_CONTENT_PER_READ | 39 | 45 | G | Mean percentage of GC bases |
| STD_GC_CONTENT_PER_READ | -1 | 15 | G | Std dev of Percentage GC |
| MEAN_BASE_QUAL_PER_POSITION | 30 | 100 | Q | Mean of mean base calling quality |
| STD_BASE_QUAL_PER_POSITION | -1 | 6 | Q | Std dev of mean base calling quality |
| MEAN_N_PER_POSITION | -1 | 10 | N | Mean Percentage N |
| STD_N_PER_POSITION | -1 | 10 | N | Std dev of Percentage N |
| MEAN_A_PER_POSITION | 25 | 35 | B | Mean Percentage A |
| STD_A_PER_POSITION | -1 | 10 | B | Std dev of Percentage A |
| MEAN_C_PER_POSITION | 15.5 | 25 | B | Mean Percentage C |
| STD_C_PER_POSITION | -1 | 10 | B | Std dev of Percentage C |
| MEAN_G_PER_POSITION | 17 | 24 | B | Mean Percentage G |
| STD_G_PER_POSITION | -1 | 10 | B | Std dev of Percentage G |
| MEAN_T_PER_POSITION | 25 | 33 | B | Mean Percentage T |
| STD_T_PER_POSITION | -1 | 10 | B | Std dev of Percentage T |
| 32_MER_ERROR_RATE | | | | Estimated 32-mer error rate |
| ADAPTER_8_MERS | -1 | 5 | A | Percentage of Universal adapter 8-mers |
| MARKED_DUPLICATE | -1 | 60 | D | Percentage marked as duplicate |
| UNMAPPED | -1 | 20 | U | Percentage unmapped reads |
| BOTH_UNMAPPED | -1 | 30 | U | Percentage both reads in pair unmapped |
| FIRST_UNMAPPED | -1 | 30 | U | Percentage only first unmapped in pair |
| SECOND_UNMAPPED | -1 | 30 | U | Percentage only second unmapped in pair |
| PROPER_PAIRS | | | | Percentage proper pairs |
| PROPER_PAIRS_AUTOSOME | 95 | 1000 | P | Percentage proper pairs autosome |
| FF_RR_PAIRS | -1 | 0.1 | o | Percentage FF/RR oriented pairs |
| MEAN_COVERAGE | 0.1 | 100000 | C | Mean coverage |
| STD_COVERAGE | -1 | 100000 | C | Std dev of coverage |
| MEAN_INSERT_SIZE | -1 | 10000 | I | Mean insert size |
| STD_INSERT_SIZE | | | | Std dev of insert size |
| ADAPTER_INSERT_SIZE | -1 | 20 | A | Percent insert size < read length |
| MAPPING_QUAL_60 | | | | Percentage reads with mapping quality <60 |
| MAPPING_QUAL_40 | | | | Percentage reads with mapping quality <40 |
| MAPPING_QUAL_20 | | | | Percentage reads with mapping quality <20 |
| MEAN_MISMATCHES | -1 | 5 | m | Mean mismatches per read pair |
| MEAN_DELETIONS | | | | Mean deletions per read pair |
| MEAN_INSERTIONS | | | | Mean insertions per read pair |
| NZ_DELETIONS | -1 | 0.1 | d | Fraction or reads that have a deletion |
| NZ_INSERTIONS | -1 | 0.1 | I | Fraction of reads that have an insertion |
| CLIPPED_5_PRIME | -1 | 6 | c | Percentage of reads clipped at 5'-end |
| CLIPPED_3_PRIME | -1 | 30 | c | Percentage of reads clipped at 3'-end |
| C>A | 0.3 | 0.7 | O | C>A triplet conversion rate |
| G>A | 0.4 | 0.6 | O | G>A triplet conversion rate |
| T>A | 0.3 | 0.7 | O | T>A triplet conversion rate |
| A>C | 0.3 | 0.7 | O | A>C triplet conversion rate |
| G>C | 0.3 | 0.7 | O | G>C triplet conversion rate |
| T>C | 0.3 | 0.7 | O | T>C triplet conversion rate |

Table S20. Metrics collected for each lane by bamqc_summary.
If any flag is raised, the lane is excluded from the merge process. The values, per read group, are collected in the file .bamqc_summary.

### Supplementary Notes

#### Supplementary Note 1: WGS data quality specification.

Sequencing was performed at the two sequencing providers, deCODE genetics and the Wellcome Sanger Institute, according to the specifications set forth in the material transfer agreement for UKB Access application nr. 52293 – Summarized as follows:

| QC parameter | Sample level | Batch level |
|---|---|---|
| Sequencer type | Illumina NovaSeq6000 or better with standard 151 base, paired-end chemistry | |
| Sequencing library | PCR-free, uniquely dual-indexed in multiplexed pools | |
| Read-length | >100bp | |
| Proper-pairs | % of mapped read-pairs from the same DNA fragment with appropriate orientation and separation:<br>≥95% PASS<br><95% FAIL | |
| Coverage | % of autosome covered ≥15x:<br>≥95% PASS<br><95% FAIL | The mean sample genome coverage across the monthly sequencing batch is expected to be approximately 30X across the genome with a minimum coverage of 26X. |
| Contamination level 1<br>(Freemix) | Freemix sample contamination level as measured by VerifyBamID[13]:<br>≥5% FAIL<br>>1% and <5% further analyzed with Read_haps[14]<br><1% PASS | ≤4 samples per 96 sample sequencing plate<br>≤1% per monthly sequencing batch |
| Contamination level 2<br>(Read_haps) | For samples with Freemix values 1-5%, contamination is verified by Read_haps | |
| Sample Identity Concordance | Discordance at non-reference genotypes ≥2% FAIL<br><2% PASS | Sample identity concordance failures within each monthly sequencing batch must be <0.05% |
| Monthly seq batch overall failure rate | | Repeat Sample requests are no more than 1% of the monthly sequencing batch |

All calculations of data quantity (yield) and coverage must exclude duplicate reads, adaptors, overlapping bases from reads from the same fragment, soft-clipped bases

#### Supplementary Note 2: Whole genome sequencing

DNA samples were selected by UK Biobank using its picking algorithm which ensures pseudo-randomisation of recruitment centres and collection times across batches, to avoid potential batch effects and shipped on dry-ice to the sequencing centers at Wellcome Sanger Institute (Sanger) in Cambridgeshire, UK (WSI) and deCODE genetics in Reykjavik, Iceland (deCODE). The two institutes then followed commensurate protocols, with one protocol at deCODE and two protocols at Sanger; Sanger Vanguard and Sanger Main.

##### deCODE protocol

The samples were in 70 µL aliquots in Fluid-X 0.3 mL, externally threaded 2D barcoded tubes in 96-well racks with linear barcodes (Brooks Life Sciences) at a normalized, target DNA concentration of 12 ng/µL in 1x TE buffer (10 mM Tris-HCl, 1.0mM EDTA, pH 8.0). Upon arrival, samples/plates were registered in the respective Laboratory Information Management System (LIMS) and stored until use at -20 °C. DNA concentration was confirmed by UV/VIS spectrophotometry (Trinean DropSense system or equivalent). Sequencing libraries were prepared using the NEBNext Ultra™ II PCR-free kit (New England Biolabs). In short, 500 ng of genomic DNA was fragmented to a mean target size of 450-500 bp using high frequency Adaptive Focused Acoustics Technology (AFA) from Covaris Inc (LE220plus instruments and 96-well TPX-AFA plates). End repair and A-tailing was performed in a single step followed by ligation of unique dual indexed sequencing adaptors (IDT for Illumina) and two rounds of SPRI-bead purification (0.6X) using an automatic 96/8-channel liquid handler (Hamilton Microlab STAR and Tecan Freedom EVO). Quality (concentration and insert size) of sequencing libraries was determined using the LabChip GX (96-samples) instrument (Perkin Elmer). Sequencing libraries were pooled appropriately using automatic 8-channel liquid handlers and sequenced using Illumina´s NovaSeq6000 instruments. Paired-end sequencing on the S4 flowcell (v1.0 chemistry) was performed with a read length of 2x151 cycles of incorporation and imaging, in addition to 2*8 index cycles to a mean coverage of at least 26X per sample. Real-time analysis (RTA) involved conversion of image data to base-calling in real-time. All steps in the workflow were monitored using the in- LIMS with barcode tracking of all samples/plates and reagents.

##### Sanger Vanguard and Main Protocol

Genomic DNA samples were received at WSI in 0.3ml externally threaded 2D barcoded FluidX tubes, held in 96-well SBS racks (Azenta Life Sciences). All samples were scanned into an in-house LIMS tracking system upon receipt and stored at -20°C. Prior to processing, samples were subjected to plate-based gravimetric assessment using a PJ-3000 laboratory balance (Mettler Toledo). To ensure sample homogeneity prior to measurement, samples were heated and agitated at 45°C,100rpm for 20 minutes in a SI500 orbital incubator (Stuart Scientific). Sample racks were subsequently secured in a DVX-2500 multi-tube vortexer (VWR) and mixed at 1400rpm for 10 minutes. Samples were quantified in triplicate using the AccuClear Ultra High Sensitivity dsDNA Quantitation kit (Biotium).

Assay setup was performed on a Mosquito LV (SPT Labtech) and Agilent Bravo NGS workstation, fluorescence was measured on a FLUOstar Omega microplate reader (BMG Labtech).

To generate PCR free libraries, genomic DNA was sheared to an average fragment size of 450bp using a LE220 focused ultrasonicator (Covaris). Library construction (end repair, A-tailing and adapter ligation) was performed using an NEBNext Ultra II custom kit (New England Biolabs) on a Bravo NGS workstation (Agilent Technologies). Samples were tagged using IDT for illumina TruSeq UD Indexes during ligation. Following an AMPure XP (Beckman Coulter) purification and size selection workflow, libraries were quantified by qPCR on a Roche LightCycler 480 using a custom KAPA kit (Roche Life Science). Equimolar pools were created on a Biomek NX-8 liquid handler (Beckman Coulter), and sequenced on the illumina NovaSeq 6000 platform using S4 flow cells and 150bp paired-end reads. Samples falling below the coverage threshold underwent top-up sequencing using the Xp workflow on S4 flow cells. Top-up data was merged with original cram files and re-processed through the standard analysis pipeline. End-to-end sample traceability was supported by the use of an in-house LIMS.

#### Supplementary Note 3: Sequence processing pipeline

Three commensurate sequence processing pipelines were developed, deCODE, Sanger Main and Vanguard.
Although different pipelines were used as a consequence of the different project stages and service providers, the final allocation of samples to Vanguard or Main Phase does not reflect the particular pipeline used.

##### deCODE pipeline

The deCODE pipeline (Fig. S12, Fig. S13) for UKB consists of the following steps. An automated pipeline monitors the data coming off the sequencers and starts processing the data when the sequence run folder is ready. The steps taken are:

1. bcl2fastq is run on the sequencer run folder to demultiplex the data and convert each (lane,index) combination into fastq pairs. A checksum is generated for each fastq pair and stored for future reference. The reads in the fastq files are counted and compared against the expected counts coming from the sequencer. The Undetermined read files are inspected, looking for reads that haven't been accounted for.
2. Each pair of fastq files is processed to create a CRAM file. The steps are
    a. Align against GRCh38
    b. Fix mate pair information
    c. Mark duplicates.
    d. Sort in genomic order
    e. calculate checksum and compare with fastq checksum. Failure if they don't match and process is rerun
3. CRAM file is compared with chip genotypes for same sample. Result reported back to the lab. Failure if mismatch rate >2% (potential sample error)
4. QC stats are collected and thresholds applied (Supplementary Fig. 14). Results are reported back to the lab and CRAM is failed if it doesn't pass all quality parameter thresholds. Failed lanes are archived and not used in further processing.

5. A merge process monitors the (lane,index) data and merges the data when it is likely that sufficient data have been collected for a sample. The merge process injects all the necessary header information into the file making it ready for export to UKB.
6. When the file has been created, a checksum is generated for each read group and compared with the corresponding checksums for the fastq files. Failure if they don't match and the merge process is rerun.
7. The merged CRAM file is archived and the upstream data are marked for deletion.
8. Variant calling is performed on the CRAM file and the result is prepared for export to UKB. This includes the production of the BQSR[15] table as well as a gVCF file.
9. QC stats for the merged file are collected and thresholds applied. Results are reported back to the lab.
    a. If the file fails on quantity only, the file is held, the lab initiates a top-up run which is processed as described above and upon completion is merged with the held CRAM file into a new merged CRAM file. That new merged CRAM file is then processed again as described above
    b. If the file fails on other quality parameters, the file is failed and the sample is flagged in the lab. The lab must decide the appropriate action (abandon sample, request a new library)
10. The merged CRAM file, along with variant calling and auxiliary data are sent to UK Biobank

#### Pipeline details

##### *Alignment*

Each read group is aligned to GRCh38 reference (GRCh38 reference with alt contigs plus additional decoy contigs and HLA genes) with bwa mem (v0.7.17)[4] using parameters '-K 100000000 -Y -t 24'. To add MC and MQ tags, samblaster[26] (v0.1.24) is used with parameters '-a --addMateTags'. Duplicates are marked using Picard MarkDuplicates (v2.20.3) with parameters "ASSUME_SORT_ORDER=queryname READ_NAME_REGEX='[a-zA-Z0-9-]+:[0-9]+:[a-zA-Z0-9]+:[0-9]:([0-9]+):([0-9]+):([0-9]+)'", then the results are coordinate sorted using samtools[2] (v1.9).

##### *Merging*

Internal thresholds are set for total sequence yield and read count, GC fraction (first and second read in pair) and bias compared to reference, flagging of base conversions in sample preparation, where certain trinucleotides are more commonly observed in sequencing than their reverse complement, flagging of base conversions in sample preparation, where certain trinucleotides are more commonly observed in sequencing than their reverse complement, percentage aligned library read pairs, library insert fragment size distribution, sequencing adapter contamination level, sequence run base call quality values, genotype concordance rate against supplied genome-wide genotype data supplied by UKB for each participant sample, sequence error rate, sequence contamination rate and genome coverage. Read group bam files are assessed for these parameters and those that pass all the thresholds are merged using samtools[2] merge (v1.9) and converted to CRAM format.

##### *Single sample variant calling*

A base quality recalibration table is created using GATK BaseRecalibrator (v4.0.12) with known sites files dbSNP138, Mills and 1000G gold standard indels, and known indels from GATK resource bundle and parameters "--preserve-qscores-less-than 6 -L chr1 .. -L chr22". For each chromosome in chr1 .. chr22, chrX, chrY, the resulting base recalibration table is applied using GATK ApplyBQSR (v4.0.12) with parameters "--preserve-qscores-less-than 6 --static-quantized-quals 10 --static-quantized-quals 20 --static-quantized-quals 30 --create-output-bam-index" and then variants are called using GATK[15] HaplotypeCaller (v4.0.12) with parameters "-ERC GVCF". The resulting 24 chromosome g.vcf files are then combined using Picard[15] MergeVcfs (v2.20.3).

*Quality assessment reports*

Reports (Supplementary Table 1) to assess the data quality are created using the following programs (in the steps Lane QC, QCPreview and QCStats):

- BamQC (v1.0.0) run on each lane before merge (Supplementary Table 2).
- samtools[2] stats (v1.9) using parameters "-d -p" , i.e. excluding duplicates and overlapping basepairs
- Picard CollectWGSMetrics (v2.20.3) is run with parameters "USE_FAST_ALGORITHM=True MINIMUM_BASE_QUALITY=0 MINIMUM_MAPPING_QUALITY=0 COVERAGE_CAP=1000" once for whole genome, once for autosomes only
- Genotypes are called from .g.vcf files using GATK GenotypeGVCFs (v4.0.12)
- Sample contamination is assessed by running verifyBamId[13] (v1.1.3) with parameters "--ignoreRG --chip-none --free-full --maxDepth 100 --precise" using 1000G phase 3 autosomal SNPs with European MAF > 0.01
- Sample contamination is accessed again using read_haps[14] "-q 30 -mq 30 -c 1 -w 1000"
- Genetic sex is determined using a set of some 100 000 chrX SNPs from gnomad with Non-Finnish European MAF > 0.2. For each variant, the genotype is called using GATK GenotypeGVCFs. Then the ratio of observed to expected heterozygosity assuming diploidy is computed. If ratio > 0.7 the sample is called female, if ratio < 0.3 the sample is called male, otherwise undetermined. Implemented using in-house script gvcf_sexcheck.py
- Picard[15] Genotypeconcordance (v2.20.3) is run with parameter "MIN_GQ=30" to determine concordance with genotypes for quality variants from a chip array.

#### Vanguard Pipeline

##### Sanger Vanguard Pipeline

The pipeline (Supplementary Fig. 5, Supplementary Fig. 6) created for UKB Vanguard at the Sanger consisted of the following steps. An automated pipeline monitors the data coming off the sequencers and starts processing the data once the sequencing run has completed. bambi (v0.11.1, 0.11.2, 0.12.0, 0.12.1) was used to demultiplex the data from the run folder and convert it to CRAM format. biobambam223 bamseqchecksum (v2.0.79) was used to generate a read count and checksum of the readnames and sequencing data to check for data consistency at the start and end of the processing within the Sanger. Demultiplexed sequencing data was merged per sample per run where relevant and QC metrics were generated and manually reviewed to ensure the data per sample met contracted criteria. Sample data meeting criteria was sent to Velsera Seven Bridges (SB) in CRAM format via a cloud bucket.

##### UK Biobank Vanguard Pipeline

All samples (44800) were processed on the EU deployment of the Velsera Seven Bridges Platform (SB Platform), a cloud-based research ecosystem that provides tools and infrastructure for orchestrating multi-modal data management, accessing multi-cloud compute resources and executing bioinformatics workflows at scale.

Data was received from Sanger in CRAM format via Cloud Storage Buckets. The Connect Cloud Storage feature was used to mount storage buckets directly to the SB Platform. This allowed for the data to be co-localized with workflows deployed on Cloud compute instances, in order to minimize data transfer steps and optimize analyses. Workflows were implemented in Common Workflow Language[27] (CWL, sbg:draft2 version) [REF]. Data processing was orchestrated programmatically with sevenbridges-python application programming interface (API) scripts.

##### *Whole Genome Sequencing Analysis Workflow with BWA, GATK, and Manta*

Whole Genome Sequencing Analysis Workflow with BWA, GATK, and Manta is based on the Broad Institute Best Practices workflows[3] and the principles of functional equivalence[2]. The workflow was used to re-align CRAM files to GRCh38 with bwa-mem[4] (0.7.17), call SNPs and small indels with GATK tools (4.0.12.0)[3], call structural variants with Manta[28] (1.4.0) and collect QC metrics with FastQC[29] (0.11.5), Picard tools[3] (2.18.26), VerifyBamID[30] (1.1.3) and SnpEff[11] (4.3k). Input CRAM files were converted to FASTQ format for downstream processing with biobambam2[31] bamtofastq (v2.0.87) and the quality of raw sequencing reads was assessed with FastQC. The reference genome version used for alignment was GRCh38 with the Epstein–Barr virus sequence, alternative (alt) contigs, decoy contigs, and HLA genes included. After alignment with bwa-mem, duplicates were marked with Picard MarkDuplicates and alignment files were coordinate-sorted and indexed (sambamba[32] sort, v0.5.9) before generating the final CRAM files and associated indices (samtools[33] v1.9, tabix[34] v0.2.6 and md5sum). The contents of the input and final CRAM output files were spot-checked with biobambam2 bamseqchksum as part of the analysis QC process.
Base quality score recalibration steps included GATK BaseRecalibrator and GATK ApplyBQSR. VerifyBamID was used to estimate cross-sample contamination, whereas alignment metrics were calculated with Picard CollectAlignmentSummaryMetrics and Picard CollectWgsMetrics tools. Germline SNPs and insertions/deletions were identified using GATK HaplotypeCaller, output single sample gVCF files were compressed and indexed using tabix tools, and corresponding MD5 checksums were generated. Manta was used to identify larger structural variations and the quality of variant calls was evaluated with Picard CollectVariantCallingMetrics and SnpEff tools. Please see https://github.com/UKBseq500k-methods for a full list of all tools and command line parameters used in the workflow.

Additional QC metrics were collected with samtools stats (v1.9) and the following exclusion read filter flags to match the practices of the sequencing provider: SECONDARY/SUPPLEMENTARY, SECONDARY/SUPPLEMENTARY/DUPLICATE,and SECONDARY/SUPPLEMENTARY/DUPLICATE/QCFAIL.

Generated data products were initially exported to Cloud buckets for archived storage and later in the project were exported to EMBL-EBI's data storage space.

##### *Genotype Concordance Workflow*

Genotype Concordance Workflow was used to evaluate concordance between WGS, WES and variants identified using array genotyping by determining NRD (non-reference discordance <2%) values using bcftools[26] stats (v1.9) over all SNP and indel sites shared with the array data (or exome data in genome-exome comparisons). A complete list of workflow parameters and tool versions is reported in https://github.com/UKBseq500k-methods.

##### *UK Biobank Array Data Preparation*

For genotype concordance comparisons, UK Biobank array data was lifted over to GRCh38 coordinates. Marker QC file (ukb_snp_qc.txt) was downloaded from the UK Biobank data showcase and used to extract A1 and A2 alleles (used to set reference alleles during conversion). PLINK[35] 1.9 recode command was used to convert the file sets for individual chromosomes. For this purpose, chromosome, position, A1 (ref) and A2 (alt) alleles were pulled from the UK Biobank array marker QC file (ukb_snp_qc.txt) and transformed to a VCF file format with awk. Numerically coded chromosomes 23, 24, and 26 were renamed to X, Y, MT in the VCF version of the marker QC file (and chr X, chrY and chrM in the output files). ChrXY data were omitted from processing. BIM, BED and FAM files were supplied separately to Plink 1.9 due to the different file names. Sample columns were named using individual IDs, allele order was kept (--keep-allele-order) and all alleles were set to the A2 allele, with reference alleles pulled from the transformed marker QC file. Plink-converted VCFs were lifted over to GRCh38 coordinates using CrossMap[36] 0.2.7. The GRCh38 VCFs were coordinate sorted (vcftools sort -c), bgzip-compressed, and tabix-indexed before being used in the concordance checks. In total, 1896 variants remained unmapped after the lift-over and were omitted from further analysis. Manual inspection of the unmapped variants and VEP rs ID mapping indicated that for most of these variants the reference allele differed between GRCh37 and GRCh38, whereas the rest could not be mapped or rs IDs were not associated with any GRCh38 coordinates.

##### Sanger Main Pipeline

The pipeline (Supplementary Fig. 7, Supplementary Fig. 8) created for UKB Main Phase at the Sanger consisted of the following steps. An automated pipeline monitors the data coming off the sequencers and starts processing the data once the sequencing run has completed. bambi (v0.12.2, 0.13.1, 0.14.0) was used to demultiplex the data from the run folder and convert it to CRAM format. biobambam2[23] bamseqchecksum (v2.0.79) was used to generate a read count and checksum of the readnames and sequencing data to check for data consistency at the start and end of the processing within the Sanger. Demultiplexed sequencing data was merged per sample per run where relevant and minimal QC metrics were generated. Metrics were auto reviewed and data was sent to SB in CRAM format via a cloud bucket unless it showed evidence of instrument based issues.

WGS samples were processed on the EU deployment of the SB Platform.

Data was received from Sanger in CRAM format via Cloud Storage Buckets. The Connect Cloud Storage feature was used to mount storage buckets directly to the Platform. This allowed for the data to be co-localized with workflows deployed on Google Cloud compute instances, in order to minimize data transfer steps and optimize analyses. Workflows were implemented in CWL[27] (sbg:draft2 version). Data processing was orchestrated programmatically with sevenbridges-python application programming interface (API) scripts and Seven Bridges RHEO automation code packages

Data processing was split into three phases so that only samples successfully passing analysis quality control criteria for a phase would advance to the next stage of processing. The first phase verified the input CRAM data integrity (CRAM Check Phase 1 CWL workflow), by checking MD5 sums for delivered files against the provided manifest and verifying the format of the data files with samtools view (v1.9). The second phase focused on data alignment and BAM processing steps (WGS Phase 2 CWL workflow). To reach the third phase, which included BQSR and small variant calling (WGS Phase 3 CWL workflow), a sample had to fulfill the following criteria: FREEMIX < 1% or (1% <= FREEMIX < 5% and read_haps-double_error_fraction < 0.2%), at least 95% of the autosomes covered to >= 15X (excluding duplicate reads, adaptors, overlapping bases from reads from the same fragment, and soft clipped bases), proportion of mapped read-pairs with appropriate orientation and separation > 95% and minimum read length > 100 bp. If a sample had insufficient coverage, but satisfied all other quality criteria, the BAM file was stored, merged with alignments data from subsequent "top-up" sequencing runs of the same sample and re-evaluated. During third phase processing, sample identity is checked against a subset of SNPs from the UK Biobank array genotype data (NRD < 2%).

##### *CRAM Check Phase 1 workflow*

This workflow verifies the input data integrity with samtool view, as shown below

| **Tool (version)** | **Parameter** | **Description** |
|---|---|---|
| Samtools View (1.9) | samtools view -u --reference Homo_sapiens.GRCh38_15_plus_hs38d1.fa input.cram \| samtools view -c | This command line is used to check the format of the input CRAM. Success codes of piped processes are evaluated via a bash wrapper around the command given. |

##### *WGS Phase 2 workflow*

Input CRAM files were converted to FASTQ format with biobambam2 bamtofastq (v2.0.144). The raw read-group level files were assessed with FastQC (v0.11.7) and mapped to GRCh38 reference genome, inclusive of the Epstein–Barr virus sequence, alternative (alt) contigs, decoy contigs, and HLA genes, with BWA-MEM (v0.7.17). After duplicate marking (Picard MarkDuplicates v2.18.26), the BAM files were coordinate sorted with sambamba (v0.5.9), converted to CRAM format and indexed (samtools v1.9). VerifyBamID (v1.1.3) is used to estimate cross-sample contamination, a custom script based on samtools idxstats (v1.9) is used to estimate genetic sex and additional QC metrics are collected with Picard CollectSequencingArtifactMetrics (v2.18.26) and samtools stats (v1.9). Biobambam2 bamseqchksum (v2.0.144) was used to verify the contents of the final CRAM output files. Please see https://github.com/UKBseq500k-methods for a full list of workflow steps and command line parameters.

##### *WGS Phase 3 workflow*

Phase 3 processing included base quality score recalibration (GATK BaseRecalibrator and GATK ApplyBQSR 4.0.12.0) and germline SNP and small insertions/deletions variant calling with GATK HaplotypeCaller (4.0.12.0). Output single sample gVCF files were compressed and indexed with tabix tools (v.0.2.6). During this phase of processing, samples with the

FREEMIX contamination value >1% but <5% were analyzed with read_haps[14] (commit g763b74e) and kept if the read_haps-double_error_fraction < 0.2%. Sample identity was also verified against a subset of UK Biobank array data SNP markers with bcftools stats (1.9).

#### Supplementary Note 4: Websites:

##### GraphTyper
https://github.com/DecodeGenetics/graphtyper

##### GATK
- *Resource bundle gs://genomics-public-data/resources/broad/hg38/v0*
- *Data processing https://github.com/gatk-workflows/gatk4-data-processing*
- *Germline calling https://github.com/gatk-workflows/gatk4-germline-snps-indels*

##### Svimmer
https://github.com/DecodeGenetics/svimmer

##### Dipcall
https://github.com/lh3/dipcall

##### RTG Tools
https://github.com/RealTimeGenomics/rtg-tools

##### bcl2fastq
https://support.illumina.com/sequencing/sequencing_software/bcl2fastq-conversion-software.html

##### Samtools
http://www.htslib.org/

##### samblaster
https://github.com/GregoryFaust/samblaster

##### BamQC
https://github.com/DecodeGenetics/BamQC

##### bambi
https://github.com/wtsi-npg/bambi

##### minimap2
https://github.com/lh3/minimap2

##### GIAB WGS samples
- *HG001 https://ftp-trace.ncbi.nlm.nih.gov/ReferenceSamples/giab/data/NA12878/NIST_NA12878_HG001_HiSeq_300x/NHGRI_Illumina300X_novoalign_bams/HG001.GRCh38_full_plus_hs38d1_analysis_set_minus_alts.300x.bam*
- *HG002 https://ftp-trace.ncbi.nlm.nih.gov/ReferenceSamples/giab/data/AshkenazimTrio/HG002_NA24385_son/NIST_HiSeq_HG002_Homogeneity-10953946/NHGRI_Illumina300X_AJtrio_novoalign_bams/HG002.GRCh38.60x.1.bam*

- *HG003 https://ftp-trace.ncbi.nlm.nih.gov/ReferenceSamples/giab/data/AshkenazimTrio/HG003_NA24149_father/NIST_HiSeq_HG003_Homogeneity-12389378/NHGRI_Illumina300X_AJtrio_novoalign_bams/HG003.GRCh38.60x.1.bam*

- *HG004 https://ftp-trace.ncbi.nlm.nih.gov/ReferenceSamples/giab/data/AshkenazimTrio/HG004_NA24143_mother/NIST_HiSeq_HG004_Homogeneity-14572558/NHGRI_Illumina300X_AJtrio_novoalign_bams/HG004.GRCh38.60x.1.bam*

- *HG005 https://ftp-trace.ncbi.nlm.nih.gov/ReferenceSamples/giab/data/ChineseTrio/HG005_NA24631_son/HG005_NA24631_son_HiSeq_300x/NHGRI_Illumina300X_Chinesetrio_novoalign_bams/HG005.GRCh38_full_plus_hs38d1_analysis_set_minus_alts.300x.bam*

- *HG006 https://ftp-trace.ncbi.nlm.nih.gov/ReferenceSamples/giab/data/ChineseTrio/HG006_NA24694-huCA017E_father/NA24694_Father_HiSeq100x/NHGRI_Illumina100X_Chinesetrio_novoalign_bams/HG006.GRCh38_full_plus_hs38d1_analysis_set_minus_alts.100x.bam*

- *HG007 https://ftp-trace.ncbi.nlm.nih.gov/ReferenceSamples/giab/data/ChineseTrio/HG007_NA24695-hu38168_mother/NA24695_Mother_HiSeq100x/NHGRI_Illumina100X_Chinesetrio_novoalign_bams/HG007.GRCh38_full_plus_hs38d1_analysis_set_minus_alts.100x.bam*

##### ENSEMBL
https://m.ensembl.org/info/data/mysql.html

##### Exon capture regions
http://biobank.ndph.ox.ac.uk/ukb/ukb/auxdata/xgen_plus_spikein.b38.bed

##### UKB data showcase
https://biobank.ndph.ox.ac.uk/showcase/search.cgi

##### Velsera (formerly Seven Bridges)

- *Platform https://www.sevenbridges.com/*
- *Connect Cloud storage https://docs.sevenbridges.com/docs/connecting-cloud-storage-overview*
- *Python API https://github.com/sbg/sevenbridges-python*
- *Rheo automation https://www.sevenbridges.com/rheo/*
- *Functionally Equivalent Workflows:*
    - *https://igor.sbgenomics.com/public/apps/admin/sbg-public-data/functional-equivalence-wgs-cwl1-0*
    - *https://igor.sbgenomics.com/public/apps/admin/sbg-public-data/gatk-pre-processing-for-variant-discovery-4-2-0-0*
    - *https://igor.sbgenomics.com/public/apps/admin/sbg-public-data/gatk-generic-germline-short-variant-per-sample-calling-4-2-0-0*

##### UKB SNP array QC files
https://biobank.ctsu.ox.ac.uk/crystal/refer.cgi?id=1955
https://biobank.ctsu.ox.ac.uk/crystal/refer.cgi?id=1955

##### ClinVar

https://ftp.ncbi.nlm.nih.gov/pub/clinvar/vcf_GRCh38/clinvar_20231007.vcf.gz
